# Lineage-informative microhaplotypes for spatio-temporal surveillance of *Plasmodium vivax* malaria parasites

**DOI:** 10.1101/2023.03.13.23287179

**Authors:** Sasha V. Siegel, Roberto Amato, Hidayat Trimarsanto, Edwin Sutanto, Mariana Kleinecke, Kathryn Murie, Georgia Whitton, Aimee R. Taylor, James A. Watson, Mallika Imwong, Ashenafi Assefa, Awab Ghulam Rahim, Nguyen Hoang Chau, Tran Tinh Hien, Justin A Green, Gavin Koh, Nicholas J. White, Nicholas Day, Dominic P. Kwiatkowski, Julian C. Rayner, Ric N. Price, Sarah Auburn

## Abstract

Challenges in understanding the origin of recurrent *Plasmodium vivax* infections constrains the surveillance of antimalarial efficacy and transmission of this neglected parasite. Recurrent infections within an individual may arise from activation of dormant liver stages (relapse), blood-stage treatment failure (recrudescence) or new inoculations (reinfection). Molecular inference of familial relatedness (identity-by-descent or IBD) based on whole genome sequence data, together with analysis of the intervals between parasitaemic episodes (“time-to-event” analysis), can help resolve the probable origin of recurrences. Whole genome sequencing of predominantly low-density *P. vivax* infections is challenging, so an accurate and scalable genotyping method to determine the origins of recurrent parasitaemia would be of significant benefit. We have developed a *P. vivax* genome-wide informatics pipeline to select specific microhaplotype panels that can capture IBD within small, amplifiable segments of the genome. Using a global set of 615 *P. vivax* genomes, we derived a panel of 100 microhaplotypes, each comprising 3-10 high frequency SNPs within <200 bp sequence windows. This panel exhibits high diversity in regions of the Asia-Pacific, Latin America and the horn of Africa (median *H*_E_ = 0.70-0.81) and it captured 89% (273/307) of the polyclonal infections detected with genome-wide datasets. Using data simulations, we demonstrate lower error in estimating pairwise IBD using microhaplotypes, relative to traditional biallelic SNP barcodes. Our panel exhibited high accuracy in predicting the country of origin (median Matthew’s correlation coefficient >0.9 in 90% countries tested) and it also captured local infection outbreak and bottlenecking events. The informatics pipeline is available open-source and yields microhaplotypes that can be readily transferred to high-throughput amplicon sequencing assays for surveillance in malaria-endemic regions.

## Introduction

The malaria parasite *Plasmodium vivax* is a major public health threat affecting the poorest and most vulnerable populations in more than 49 endemic countries^1^. Over the past decade, enhanced malaria control efforts in areas outside of sub-Saharan Africa have achieved a marked decline in *P. falciparum* infections, but a relative rise in the proportion of *P. vivax* cases^1^. Several biological attributes of *P. vivax* render this species more resilient to interventions than *P. falciparum*^*2*^. *P. vivax* forms dormant liver stage parasites (hypnozoites) that can reactivate weeks to months after initial inoculation causing recurrent episodes of malaria (relapses). A single mosquito inoculation can cause multiple relapses, sustaining transmission over extended periods. Relapses are thought to cause over 60% of clinical cases^3^. Knowledge of the biology and epidemiology of *P. vivax* relapse, including the underlying reactivation mechanism(s) and host and parasite determinants, are vital for informing public health strategies to combat this parasite, but our understanding of these processes is limited^4^. A major obstacle to our increased understanding has been the difficulty of classifying recurrent *P. vivax* infections accurately. These recurrences can arise from reinfection (new mosquito inoculations), recrudescence (blood-stage treatment failures), or relapse (reactivation of dormant liver stages). Discriminating between these causes is challenging.

Accurate methods to disentangle relapse from reinfection and recrudescence events are critical to improving our understanding of the therapeutic efficacy of current treatment regimens for *P. vivax*. Accumulating reports suggest that chloroquine, the most widely used drug for treating the *P. vivax* blood stages, is failing in several endemic regions, but recrudescence (drug failure) can be confused with relapse, confounding efficacy studies^5^. Accurate diagnosis of the cause of recurrent infection is also essential to clinical studies of hypnozoiticidal (anti relapse) regimens (primaquine and tafenoquine) as this relies on distinguishing relapse from reinfection^6,7^. Reinfection dilutes observed differences in recurrence between hypnozoiticidal interventions and biases treatment effect estimates towards interventions which provide longer post-treatment prophylaxis^8^.

The ability to discriminate between recurrent *P. falciparum* infections deriving from reinfections (which are likely genetically heterologous to the initial infection given sufficient population diversity) and recrudescences (which contain homologous parasites) using genotyping at a handful of polymorphic markers, represented a crucial advance for *P. falciparum* clinical research^9^. It allowed therapeutic efficacy studies to be conducted in endemic areas without the need for concomitant detailed epidemiological assessment. However, the situation is more complex for *P. vivax* malaria since relapsing parasites can be genetically homologous or heterologous to the initial infection^10,11^. However, recent genomic studies have revealed that a proportion of paired *P. vivax* isolates (from acute and recurrent infections) that would be classified as heterologous using traditional genotyping approaches share homology in large segments of the genome, inferring familial relatedness^12-14^. In mosquito hosts capturing blood meals with mixtures of parasite genomes, the obligate meiotic stage will generate meiotic recombinants, producing sporozoites that share parents. Every natural infection deriving from more than one sporozoite may comprise mixtures of meiotic recombinants. Pairs of infections with evidence of recent identity by descent (IBD) consistent with close relationships such as siblings or half-siblings (as much as ≥50% and ≥25% IBD respectively), are more likely to have derived from the same mosquito inoculation than from different inoculations and are, therefore, more likely to reflect relapse than reinfection events. IBD therefore has significant potential to enable more accurate classification of recurrent *P. vivax* infections. IBD-based measures would also allow finer resolution of the spatial connectivity between *P. vivax* populations than is possible using classical methods such as the fixation index or phylogenetic approaches^15^. With appropriate genetic data, IBD can be used to characterise major transmission networks (foci) for targeted intervention, or the risks of infection spread between communities and across borders.

While genomic data provides the greatest information to estimate IBD, *P. vivax* patient isolates often have low parasite densities which currently precludes whole genome sequencing at large scale, even when using selective whole genome amplification methods^16^. Restricting analyses to infections with high parasite densities is not ideal as these are atypical of *P. vivax* infection and hence may not be representative of the true diversity in patient infections. High-throughput genotyping offers a more operationally feasible approach that can be applied to low volume samples such as dried blood spots and would be more readily implemented in surveillance frameworks in malaria-endemic countries^17^. However, selecting parsimonious marker sets that can capture genome wide IBD effectively is challenging^18^. To date, genotyping methods for *P. vivax* have relied on either capillary sequencing of microsatellite markers or next generation sequencing (NGS) of Single Nucleotide Polymorphisms (SNPs)^2^. In a recent *P. falciparum* study, targeted NGS of short regions comprising multiple highly variable SNPs (microhaplotypes) provided a simpler, cheaper, and higher-throughput approach than microsatellite typing, with substantially higher resolution of individual clones than with single SNPs^19^.

Using 1,816 publicly available *P. vivax* genomes, we identified microhaplotype markers that could estimate IBD across pairs of infections in diverse parasite populations. The goal was to establish a universal *P. vivax* IBD barcode to improve the interpretation of therapeutic evaluations of hypnozoiticidal and schizonticidal antimalarial drugs, elucidate the biology and epidemiology of *P. vivax* relapses, and provide NMCPs and other agencies with actionable information on parasite transmission within and across national borders. *In silico* analyses confirmed that *P. vivax* microhaplotypes can provide much higher resolution to estimate relatedness consistent with close relationships compared to biallelic SNP barcodes that are currently used.

## Results

### Microhaplotype candidate selection

From an initial set of 1,816 isolates from the MalariaGEN Pv4 data release (**Figure 1a**), we identified 615 high quality apparently monoclonal infection samples (>50% genome positions callable, *F*ws ≥0.95)^20^. This sample set was used to select variants and identify potential microhaplotype candidate regions for targeted sequencing, with the aim of identifying 200 bp regions of the *P. vivax* genome with high diversity in isolates from all geographic regions. In addition to country classifications, the samples were assigned regional population classifications derived from the MalariaGEN Pv4 project and based on genetic clustering patterns^20^. Although most parasites were from the Asia-Pacific region, the sample set exhibited broad geographic representation, spanning 17 countries and with all regional populations represented by at least 30 isolates (range n=31-151) (**Table 1, Supplementary Figure 1**).

**Figure 1.**
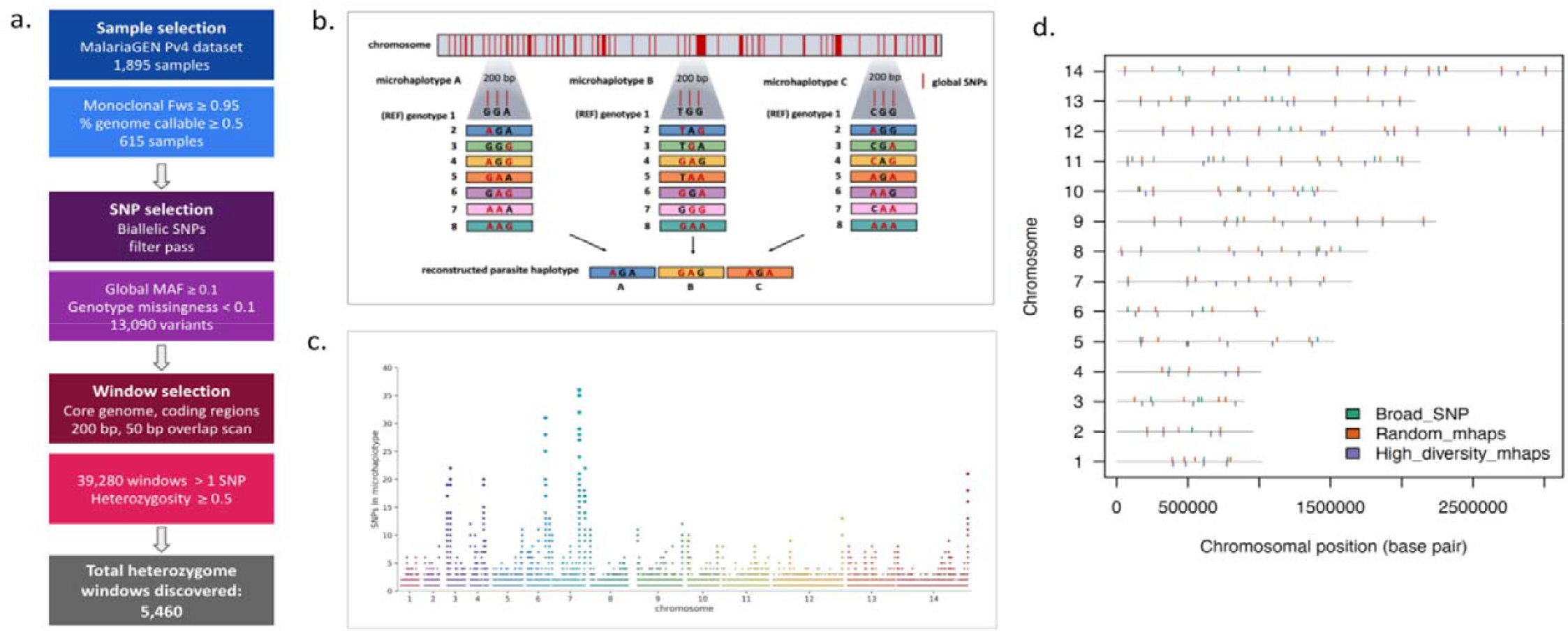
Microhaplotype discovery pipeline. Panel a) provides an overview of the marker selection process. Criteria for selecting samples, variants, and windows for potential microhaplotype candidate windows result in a total of 5,460 windows (200 bp). The MalariaGEN Pv4 dataset was filtered to use only high-quality monoclona samples (*F*ws ≥0.95) that had at least 50% of the core genome positions callable. SNP variants from this sample subset were then identified as biallelic, have low genotype missingness (<0.1), had high global minor allele frequencies (MAF ≥0.1), and FILTER = PASS in the MalariaGEN dataset, resulting in 13,090 total SNPs. The co genome was then scanned in coding regions for all 200 bp windows in which > 1 of the identified variants were found and filtered for high diversity (global heterozygosity ≥0.5). Panel b) provides a schematic representation of microhaplotypes with 3 SNPs. Microhaplotypes leverage SNP information content in small-windowed regions of the genome to provide a high-resolution reconstruction of the parasite genome. Three high-diversity SNPs in a single microhaplotype can have as many as 8 distinct combinations of alleles which, when combined with 100 microhaplotypes across the genome, results in high discriminatory power to characterize relatedness. Panel c) provides a map of the *P. vivax* heterozygome. Chromosomal distribution of all windows identified in the global set of high-quality, independent monoclonal infections with at least 1 SNP. Each point is an identified window, with the size increasing as the number of SNPs within the window increases. Potential microhaplotype regions are well distributed across the 14 chromosomes. The microhaplotypes with the highest SNP densities tend to be located at the ends of the chromosomes. Note, microhaplotypes were selected only from the accessible regions of the genome i.e., excluding highly diverse telomeric and sub-telomeric regions where sequence reads could not be mapped accurately. Panel d) illustrates the chromosome distribution of three panels evaluated for their capacity to reconstruct parasite relatedness. Two new microhaplotype panels were created from the microhaplotype discovery pipeline, named “Random mhaps” plotted in orange, and “High-diversity mhaps” plotted in purple. These two panels were selected to have 100 microhaplotype markers using windows that were well-spaced and had between 3-10 SNPS, with even distribution across all 14 chromosomes and a minimum diversity with heterozygosity ≥0.5. The markers for the Random mhaps panel were selected randomly, while the High-diversity mhaps panel were optimised to have the highest heterozygosity possible in each region. These two high-resolution panels were compared to the currently used biallelic 42-SNP panel, named “Broad” in green. Only 38 markers of this panel are considered informative and included in this representation.

**Table 1.** Geographic distribution of the sample set. The countries represented within each regional group are Ethiopia (AF), Cambodia, Thailand and Vietnam (ESEA), Malaysia (MSEA), Indonesia and Papua New Guinea (OCE), Brazil, Colombia, El Salvador, Mexico, Nicaragua and Peru (SAM), Afghanistan, India and Iran (WAS), and Thailand (WSEA). The Thai samples in ESEA and in WSEA derive from provinces east and west respectively of a previously described malaria-free corridor that runs through the centre of Thailand^42^.

| Region | No. isolates sequenced | No. high-quality independent monoclonal isolates | No. high-quality paired isolates |
| --- | --- | --- | --- |
| Africa (AF) | 47 | 34 | 0 |
| East Southeast Asia (ESEA) | 262 | 151 | 6 |
| Maritime Southeast Asia (MSEA) | 76 | 59 | 4 |
| Oceania (OCE) | 205 | 120 | 2 |
| South America (SAM) | 159 | 135 | 0 |
| West Asia (WAS) | 46 | 31 | 0 |
| West Southeast Asia (WSEA) | 127 | 85 | 2 |
| **Total** | **922** | **615** | **14** |

A total of 13,090 candidate high quality biallelic SNPs with global (pooled across regions) high minor allele frequency (MAF >0.1) and low genotype failure rate (missingness < 0.1) were identified in coding regions of the genome (excluding hypervariable regions) to minimise potential changes in primer-binding sites and missingness) (**Figure 1b)**. We then conducted a windowed scan in partially overlapping windows (200 bp windows, 50 bp scanning increments) across all identified variants of interest (39,280 windows). As illustrated in **Figure 1c**, these windows were relatively uniformly distributed across the genome. We then filtered further by heterozygosity to be above the theoretical maximum for a single biallelic SNP of 0.5. Together, these filters identified a total of 5,460 microhaplotype candidate “heterozygome windows” within the nuclear genome. Of these candidates, 100 microhaplotypes with roughly even spacing across all 14 chromosomes were chosen for further panel evaluation that each contained between 3-10 candidate SNPs, optimised for highest heterozygosity (High-diversity SNP panel: mean het = 0.82, min = 0.61, max = 0.96). A second panel of 100 randomly selected microhaplotypes (Random-SNP panel: containing any number of SNPs, heterozygosity > 0.5, evenly spaced) was also included for comparative analysis to investigate the impact of heterozygosity in panel informativeness for several use cases (**Figure 1d**). Details on the coordinates, SNPs and genes covered by the High-diversity and Random-SNP microhaplotype panels are provided in Supplementary **Table 1**. The two hypothetical microhaplotype panels (High-diversity SNP panel, Random-SNP panel) as well as a previously developed biallelic SNP panel (the 42-SNP Broad barcode)^21^ were then benchmarked for their ability to discriminate relatedness with IBD estimation among real and simulated infection pairs. Four SNPs within the Broad barcode had to be excluded from analyses because of high genotype failure rates (i.e., less than 5 reads covering the SNP in >10% of the available global *P. vivax* sample set), or multi-allelic status (i.e., not bi-allelic). This left 38 SNPs for evaluation; herein we refer to this panel as the 38-SNP Broad barcode.

### Effective estimation of identity by descent with microhaplotype panels

A formal evaluation of the accuracy of the three panels to estimate IBD between monoclonal *P. vivax* infections was first conducted using simulated data generated on pairs of infections across a range of relatedness (*r*) values using the *paneljudge* package^22^. All data were simulated under the model used to estimate relatedness. In each geographic region, the 95% confidence intervals (CIs) around the estimate 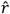 were calculated as a measure of the informativeness of a given marker panel (**Figure 2**). In all geographic regions and for all marker panels, the CI intervals were shortest when *r*=1 (identical infections), closely followed by *r*=0 (strangers), and highest when *r*=0.5 (siblings) indicating lowest accuracy in predicting sibling relationships. In all geographic regions, the CI intervals were shortest around the estimate 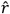 at the High-Diversity SNP microhaplotype panel followed by the Random-SNP microhaplotype panel, and the 38-SNP Broad barcode. At the High-Diversity panel, all (100%) of the estimates of *r*=0.5 had 95% CIs between 0.1 and 0.8. Whilst the Random-SNP microhaplotype panel followed similar trends to the High-diversity SNP panel in each geographic region, the Broad barcode displayed particularly large CI intervals relative to the microhaplotype panels in Maritime Southeast Asia and Oceania. The same trends in marker panel performance were observed using the root mean squared error (RMSE) of the estimates of *r* 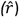 compared to the data-generating *r* (**Supplementary Figure 2**). In all regions, the High-diversity SNP microhaplotype panel exhibited consistently lower RMSE values than the Random-SNP microhaplotype and the 38-SNP Broad barcode. These results indicate that panels of ∼100 uniformly spaced microhaplotypes provide more accurate estimation of IBD than the current single SNP panel used for *P. vivax* infections; this advantage may be more pronounced in certain geographic regions, possibly reflecting the geographic representation informing SNP selection.

**Figure 2.**
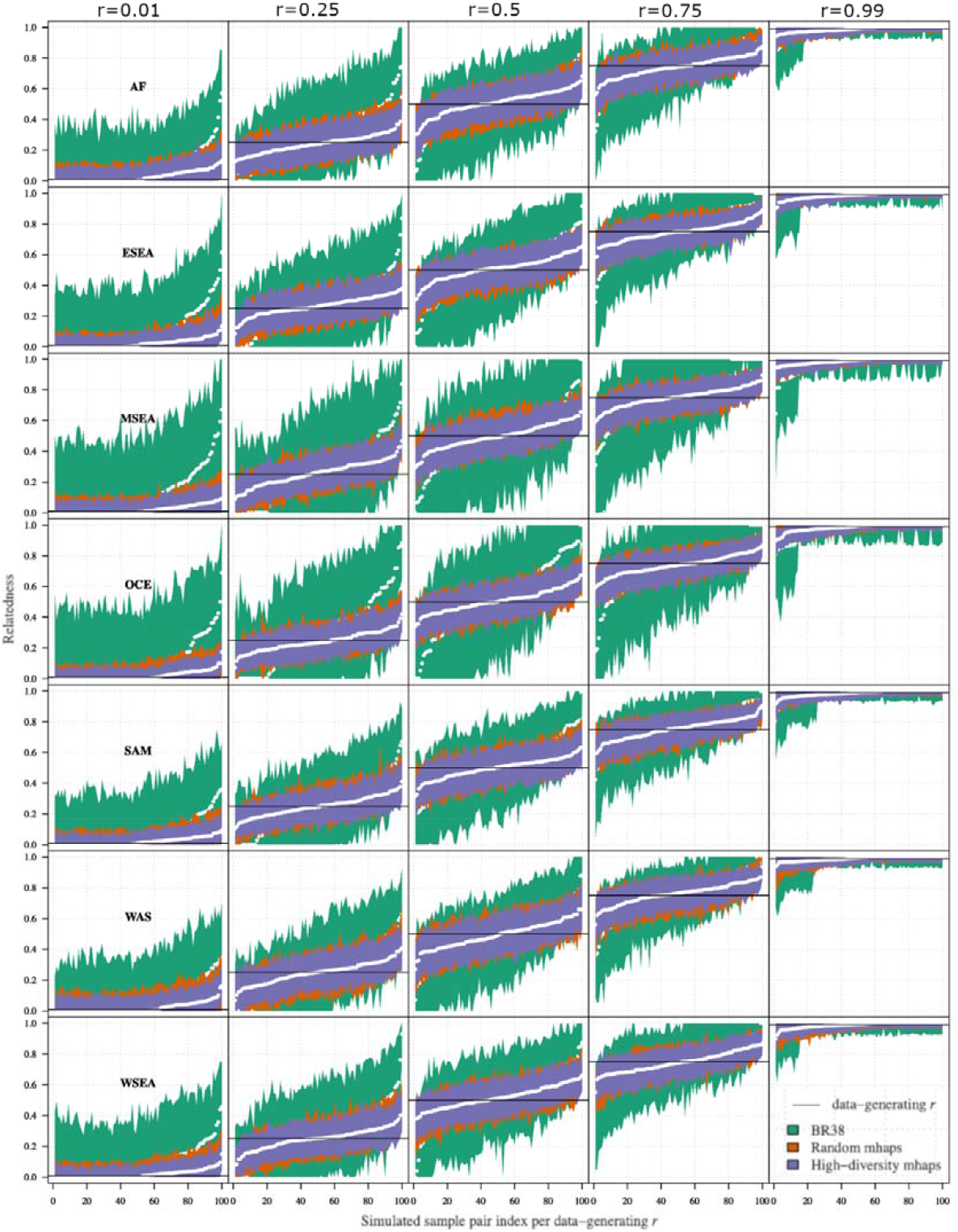
Confidence intervals around relatedness estimates based on data simulated using various data-generating relatedness parameters, r. Data are presented on 3 marker panels: High-diversity microhaplotype panel, Random-SNP microhaplotype panel, and 38 Broad barcode biallelic SNPs. Separate plots are provided for each *r* and geographic region; AF (Africa), ESEA (East Southeast Asia), MSEA (Maritime Southeast Asia), OCE (Oceania), SAM (South America), WAS (West Asia) and WSEA (West Southeast Asia).

In addition to the simulated data, the efficacy of the three panels in estimating IBD between infections was also evaluated using ‘real’ data from sequenced pairs of primary and recurrent *P. vivax* infections. These samples came from a range of clinical trials, as well as from returning travellers^23-26^. A total of 14 infection pairs satisfied the criteria for selection which required monoclonal infections with high-quality genomic data at both time points. The infection pairs originated from a range of geographic locations in the Asia-Pacific and exhibited a range of durations between the primary and recurrent episode (**Table 2**). Genome-wide IBD measures using *hmmIBD* revealed that 11 infection pairs were clones (IBD ≥0.95) reflecting potential recrudescence or relapse events, whilst 1 pair was distant relatives (0.05≤ IBD <0.25) and 2 pairs were strangers (IBD <0.05). The distant relatives and strangers reflect potential reinfection or relapse events. When the data were restricted to the 100-microhaplotype markers, the resulting *hmmIBD*-based estimates correlated highly with the genome-wide measures of IBD (rho=0.790, Spearman’s rank correlation). One pair of infections (PJ0167-C and PJ0166-C) that were classified as strangers with genomic data (IBD = 0.048) were classified as distant relatives (IBD = 0.156) with the microhaplotypes, but the other 13 pairs had concordant recurrence classifications.

**Table 2.** Genome-wide versus microhaplotype-based identity by descent (IBD) estimates in *P. vivax* infection pairs. List of sample pairs evaluated for relatedness as a fraction of the genome IBD using genome-wide and microhaplotype-based data. IBD fractions were determined using *hmmIBD* software, with microhaplotype variants analysed as biallelic SNPs. Clinical survey treatments; TQ (tafenoquine), PQ (primaquine), AS (artesunate), CQ (chloroquine), AM (artesunate-mefloquine). *Recurrence classification; clone (IBD≥0.95), close relative (0.25≤IBD<0.95), distant relative (0.05≤IBD<0.25), stranger (IBD<0.05). **Returning travellers presenting at the Royal Darwin Hospital, Darwin, Australia.

| Initial infection | Recurrent infection | Patient sampling framework | Site | Region | Year | No. days until recurrence | Genome-wide IBD fraction (*recurrence classification) | Microhaplotype IBD fraction (*recurrence classification) |
| --- | --- | --- | --- | --- | --- | --- | --- | --- |
| VVX01694 | VVX01695 | Clinical survey: TQ vs PQ<sup>23</sup> | Oddar Meanchey, Cambodia | ESEA | 2015 | 43 | 1.000 (clone) | 1.000 (clone) |
| VVX01743 | VVX01744 | Clinical survey: TQ vs PQ<sup>23</sup> | Oddar Meanchey, Cambodia | ESEA | 2015 | 169 | 1.000 (clone) | 1.000 (clone) |
| VVX01078 | VVX12129 | Clinical survey: 7- vs 14-day PQ<sup>24</sup> | Dak O, Vietnam | ESEA | 2015 | 332 | 0.997 (clone) | 0.973 (clone) |
| VVX01026 | VVX12145 | Clinical survey: 7- vs 14-day PQ<sup>24</sup> | Krong Pa, Vietnam | ESEA | 2015 | 130 | 0.070 (distant relative) | 0.110 (distant relative) |
| VVX12151 | VVX12078 | Clinical survey: 7- vs 14-day PQ<sup>24</sup> | Krong Pa, Vietnam | ESEA | 2016 | 53 | 1.000 (clone) | 1.000 (clone) |
| VVX12078 | VVX12150 | Clinical survey: 7- vs 14-day PQ<sup>24</sup> | Krong Pa, Vietnam | ESEA | 2016 | 49 | 1.000 (clone) | 1.000 (clone) |
| VVX01303 | VVX01304 | Clinical survey: TQ vs PQ<sup>23</sup> | Bangkok, Thailand | WSEA | 2014 | 61 | 0.999 (clone) | 0.977 (clone) |
| PD0607-C | PD0623-C | Clinical survey: AS vs CQ vs CQ+PQ<sup>25</sup> | Tak, Thailand | WSEA | 2001 | 136 | 1.000 (clone) | 0.977 (clone) |
| PY0023-C | PY0023-CW | Clinical survey: CQ vs AM<sup>26</sup> | Sabah, Malaysia | MSEA | 2014 | 12 | 1.000 (clone) | 0.975 (clone) |
| PY0109-CWx | PY0109-Cx | Clinical survey: CQ vs AM<sup>26</sup> | Sabah, Malaysia | MSEA | 2013 | 29 | 1.000 (clone) | 0.937 (clone) |
| PY0081-C | PY0067-C | Clinical survey: CQ vs AM<sup>26</sup> | Sabah, Malaysia | MSEA | 2014 | 29 | 1.000 (clone) | 1.000 (clone) |
| PY0082-C | PY0072-C | Clinical survey: CQ vs AM<sup>26</sup> | Sabah, Malaysia | MSEA | 2014 | 28 | 1.000 (clone) | 1.000 (clone) |
| PJ0005-C | PJ0009-C | Returning travellers** | Papua Indonesia | OCE | 2010 | 861 | 0.036 (stranger) | 0.000 (stranger) |
| PJ0167-C | PJ0166-C | Returning travellers** | Papua Indonesia | OCE | 2013 | 71 | 0.048 (stranger) | 0.156 (distant relative) |

### Microhaplotype panels can effectively capture diversity and differentiation

Further evaluation of the highest performance panel, the High-diversity SNP microhaplotype panel, was conducted to assess its utility in capturing key population genetic features. The genetic diversity of the High-diversity SNP panel was assessed for isolates from each of the 7 geographic regions using measures of heterozygosity and effective cardinality. Heterozygosities across the microhaplotypes varied, but overall diversity was high in all geographic regions (median > 0.70, i.e., >70% chance on average that two randomly selected isolates will differ at a marker) (**Table 3, Figure 3a**). The lowest diversity was observed in the horn of Africa (median heterozygosity = 0.70) and the highest in Oceania (median heterozygosity = 0.81). Similar trends occurred in effective cardinality (roughly equivalent to the number of alleles, with adjustment for minor allele frequency), with median values ranging from 3.88 in Africa to 5.18 in Oceania (**Table 3, Figure 3b**). The highest effective cardinality observed in the data set was a microhaplotype with a score of 36.9 (roughly 36 different alleles at a single microhaplotype marker) in West Southeast Asia (**Table 4**). A list of the microhaplotypes amongst the top three most diverse markers in each region is presented in Supplementary Table 2. Microhaplotype PvP01_05_v1:1369384, located in PVP01_0532400 (cysteine-rich protective antigen), is amongst the 3 most diverse markers in all 7 regions.

**Table 3.** Regional patterns of population diversity using the High-diversity microhaplotype panel. Measures were conducted on n= 615 high-quality biologically independent monoclonal samples.

| Region | Median heterozygosity (min-max) | Median effective cardinality (min-max) |
| --- | --- | --- |
| AF | 0.74 (0-0.95) | 3.88 (1-18.94) |
| ESEA | 0.77 (0.55-0.95) | 4.41 (2.22-21.28) |
| MSEA | 0.76 (0.39-0.96) | 4.16 (1.65-23.91) |
| OCE | 0.81 (0.53-0.96) | 5.18 (2.12-26.05) |
| SAM | 0.80 (0.55-0.96) | 5.07 (2.2-27.16) |
| WAS | 0.70 (0.12-0.91) | 3.36 (1.13-11.72) |
| WSEA | 0.79 (0.48-0.97) | 4.84 (1.92-36.9) |

**Table 4.**
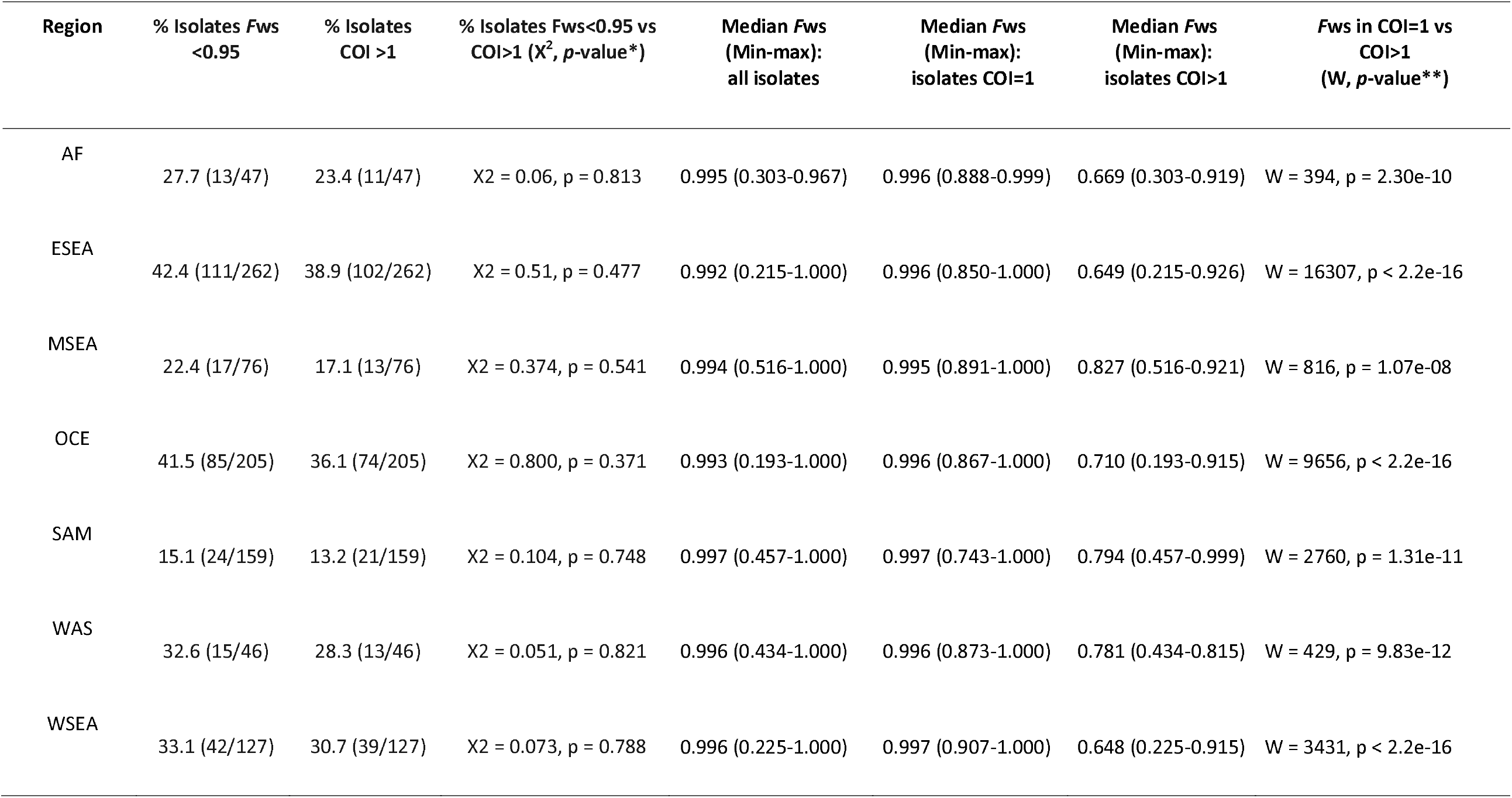
Regional patterns of within-host infection diversity. Measures were conducted on n=922 high-quality biologically independent samples. *Pearson’s Chi-squared test with Yates’ continuity correction. **1-sided Mann-Whitney U testing the hypothesis that the *F*ws in the COI=1 infection group is not larger than in the COI>1 infection group.

| Region | % Isolates *Fws* <0.95 | % Isolates COI >1 | % Isolates *Fws*<0.95 vs COI>1 ($X^2$, *p*-value*) | Median *Fws* (Min-max): all isolates | Median *Fws* (Min-max): isolates COI=1 | Median *Fws* (Min-max): isolates COI>1 | *Fws* in COI=1 vs COI>1 (*W*, *p*-value**) |
| --- | --- | --- | --- | --- | --- | --- | --- |
| AF | 27.7 (13/47) | 23.4 (11/47) | $X^2 = 0.06$, *p* = 0.813 | 0.995 (0.303-0.967) | 0.996 (0.888-0.999) | 0.669 (0.303-0.919) | *W* = 394, *p* = 2.30e-10 |
| ESEA | 42.4 (111/262) | 38.9 (102/262) | $X^2 = 0.51$, *p* = 0.477 | 0.992 (0.215-1.000) | 0.996 (0.850-1.000) | 0.649 (0.215-0.926) | *W* = 16307, *p* < 2.2e-16 |
| MSEA | 22.4 (17/76) | 17.1 (13/76) | $X^2 = 0.374$, *p* = 0.541 | 0.994 (0.516-1.000) | 0.995 (0.891-1.000) | 0.827 (0.516-0.921) | *W* = 816, *p* = 1.07e-08 |
| OCE | 41.5 (85/205) | 36.1 (74/205) | $X^2 = 0.800$, *p* = 0.371 | 0.993 (0.193-1.000) | 0.996 (0.867-1.000) | 0.710 (0.193-0.915) | *W* = 9656, *p* < 2.2e-16 |
| SAM | 15.1 (24/159) | 13.2 (21/159) | $X^2 = 0.104$, *p* = 0.748 | 0.997 (0.457-1.000) | 0.997 (0.743-1.000) | 0.794 (0.457-0.999) | *W* = 2760, *p* = 1.31e-11 |
| WAS | 32.6 (15/46) | 28.3 (13/46) | $X^2 = 0.051$, *p* = 0.821 | 0.996 (0.434-1.000) | 0.996 (0.873-1.000) | 0.781 (0.434-0.815) | *W* = 429, *p* = 9.83e-12 |
| WSEA | 33.1 (42/127) | 30.7 (39/127) | $X^2 = 0.073$, *p* = 0.788 | 0.996 (0.225-1.000) | 0.997 (0.907-1.000) | 0.648 (0.225-0.915) | *W* = 3431, *p* < 2.2e-16 |

**Figure 3.**
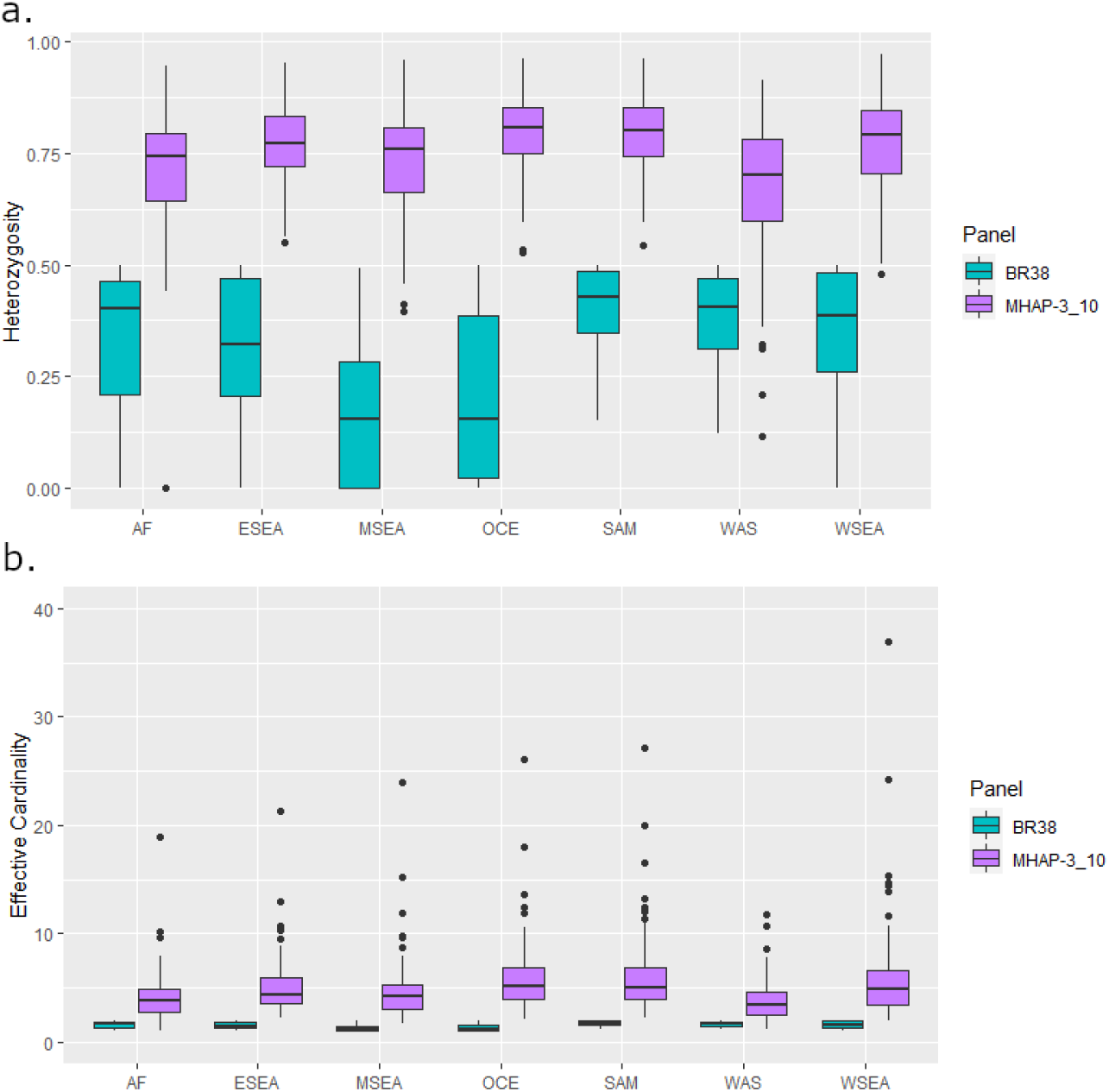
Comparative diversity between the High-diversity microhaplotype panel and the 38-SNP Broad Barcode. Panel a) presents heterozygosity measures and panel b) presents effective cardinality scores in n=615 high-quality biologically independent monoclonal samples by panel and region. Panel labels; 38-SNP Broad barcode (BR38) and High-diversity microhaplotype panel (MHAP-3_10). Regional labels; Africa (AF), East Southeast Asia (ESEA), Maritime Southeast Asia (MSEA), Oceania (OCE), South America (SAM), West Asia (WAS) and West Southeast Asia (WSEA).

Within-host diversity estimates using the High-diversity SNP microhaplotype panel were generated for each isolate using Complexity of Infection (COI) measures and correlated with the genome-wide estimates derived from the *F*_*WS*_ score. Using thresholds of COI = 1 and *F*_*WS*_ ≥ 0.95 to define a monoclonal infection, there were no significant differences in the proportion of polyclonal infections defined by the microhaplotype and genome-wide data in any geographic region (**Table 4**). Furthermore, as illustrated in **Figure 4**, the *F*_*WS*_ scores of the infections with COI=1 was significantly larger than those with COI>1 in all regions, demonstrating a strong alignment between the microhaplotype and genomic data (**Table 4**).

**Figure 4.**
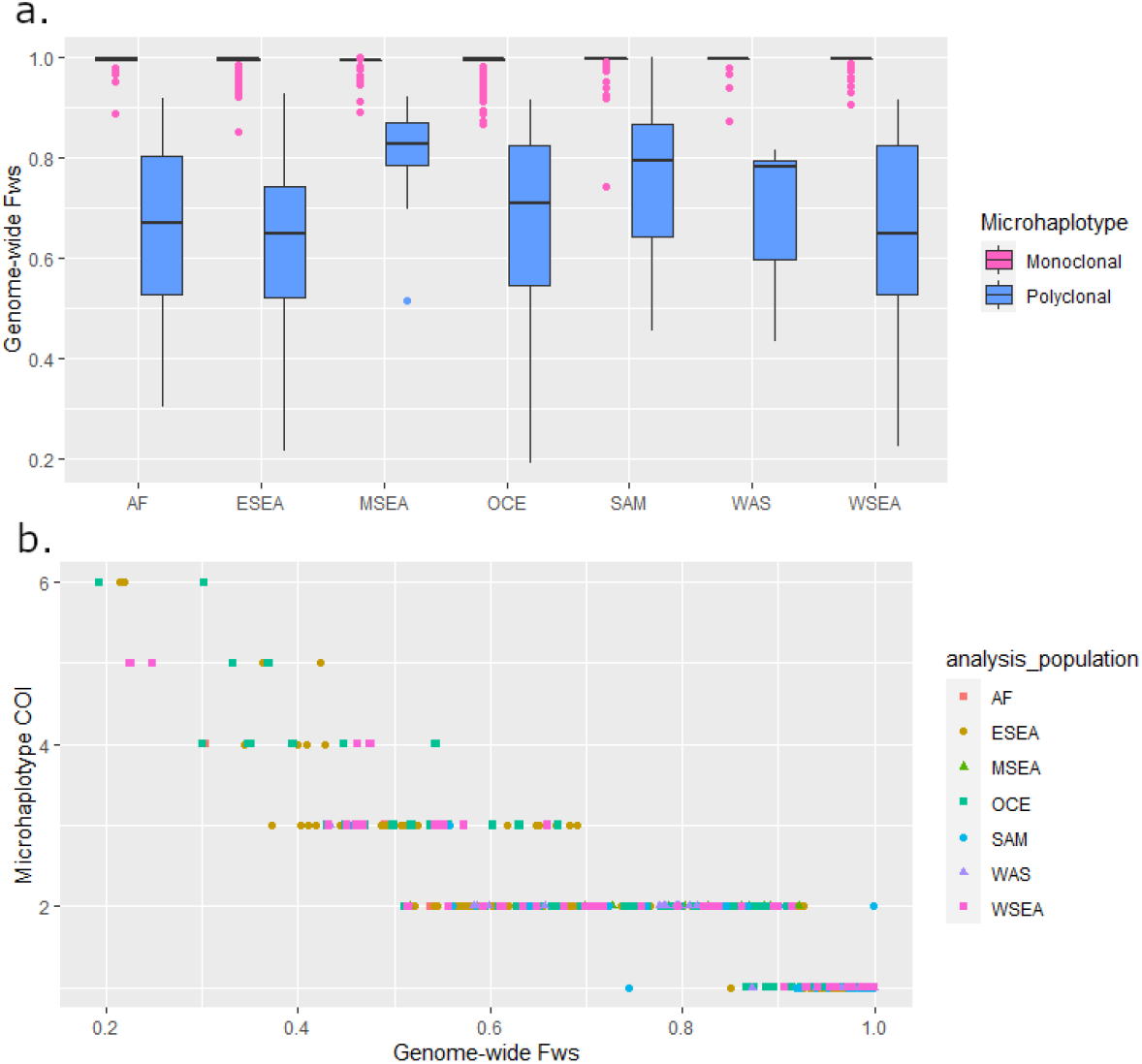
Genome-wide *F*_WS_ distribution by microhaplotype-based complexity of infection (COI). Data from n=922 high-quality biologically independent samples from Africa (AF), East Southeast Asia (ESEA), Maritime Southeast Asia (MSEA), Oceania (OCE), South America (SAM), West Asia (WAS) and West Southeast Asia (WSEA). Panel a) provides boxplots illustrating the distribution of genome wide *F*_WS_ scores in each of the monoclonal and polyclonal infection subsets as determined by THEREALMcCOIL analysis of the SNPs in the 100 microhaplotypes using the proportional function. In all geographic regions, the median genome-wide *F*_WS_ scores are closer to 1 (little to no within-host diversity) in the infections defined as monoclonal. Panel b) illustrates the correlation between genome-wide *F*_WS_ and microhaplotype-based COI estimates; a trend of decreasing COI is observed with increasing *F*_WS_ (i.e., decreasing within-host diversity).

The capacity of the High-diversity SNP microhaplotype panel to capture spatial transmission patterns including differentiation of geographically distinct populations was also assessed, using Principal Coordinate Analysis (PCoA) and IBD-based networks. The microhaplotype-based PCoA trends were consistent with spatial trends observed with genome-wide datasets^20^. Analysis of all the available *P. vivax* data demonstrated three major clusters with PC1 and PC2 representing South America, Africa and West Asia (group 1), East and West Southeast Asia (group 2), and Oceania and Maritime Southeast Asia (group 3) (**Figure 5a**). Clear differentiation of the regional groups was observed within each of the major clusters. To retain accuracy in MAF estimates, IBD-based analyses were restricted to within each of the 7 regional groupings. In all geographic regions, the microhaplotype-based estimates of pairwise IBD demonstrated a significant positive correlation with the genome-wide estimates (all *P*<0.05, **Supplementary Figure 3**). In Maritime Southeast Asia, a major clonal outbreak in Malaysia (defined K2) that we have described previously with genomic data^27^, was also distinguished clearly with the microhaplotypes (**Figure 5b**). Furthermore, more subtle sub-structure, including the resolution of previously defined K3 and K4 Malaysian sub-populations was also captured with the microhaplotypes. In other geographic regions, patterns of infection relatedness were largely consistent with genomic patterns described in previous studies (**Supplementary Figure 4**)^20^. The largest infection networks were observed in South America and Papua New Guinea. Small networks were observed in other regions, but most infections were unrelated at a minimal IBD threshold of 0.25 (consistent with half-siblings or closer). In East Southeast Asia, cross-border networks were observed between Thailand and Vietnam.

**Figure 5.**
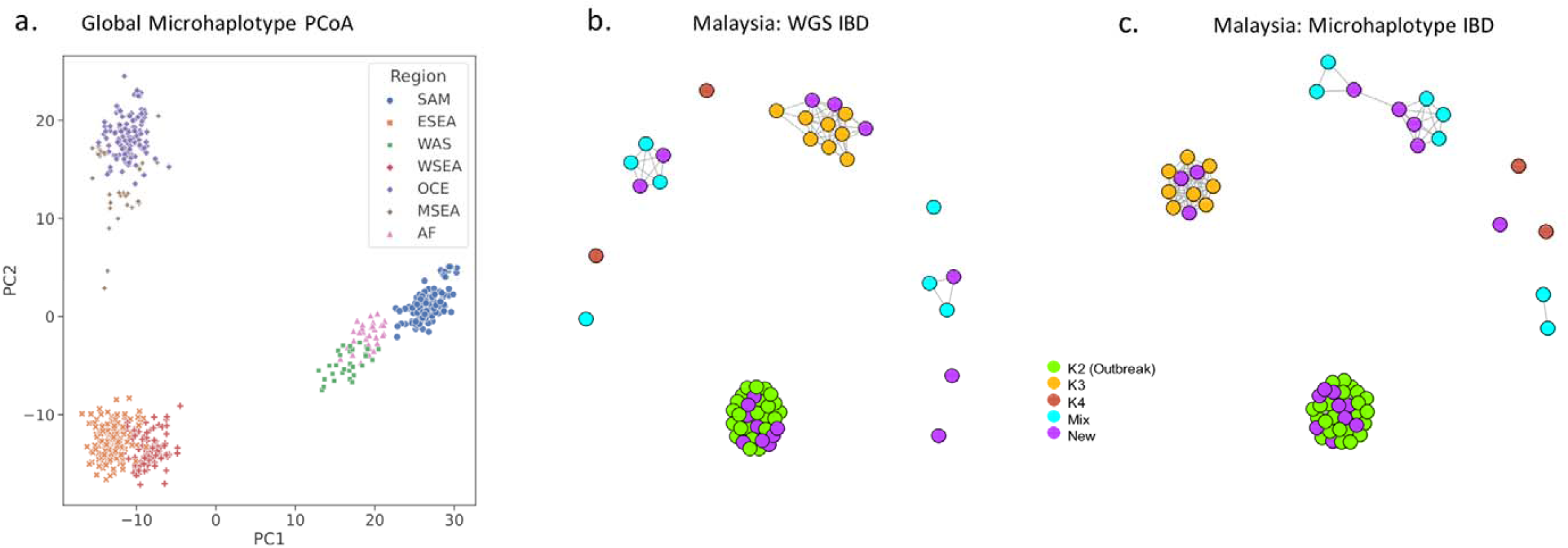
Spatial trends in *P. vivax* connectivity using microhaplotypes. Panels a) presents a PCoA plot presenting PC1 (46.4%) against PC2 (25.3%) in n= 615 high-quality biologically independent monoclonal isolates from Pv4.0 at the High-diversity SNP microhaplotype set. The combinations of PC1 and PC2 provides marked separation of all 7 regional groups. Panels b) and c) present whole genome sequencing (WGS) and microhaplotype-based IBD infection networks in Malaysia. The network plots were generated at a set of 224,612 SNPs (WGS) and the High-diversity SNP microhaplotype panel in single clone Malaysian infections (n=57) at a connectivity threshold of minimum IBD 0.5 (siblings or greater relatedness). Infections are colour-coded according to sub-structure definitions based on previously described ADMIXTURE analysis with genomic data^27^. Infections defined as “New” (purple) were not available in the previous analysis. The clustering patterns of the WGS and microhaplotype-based data are highly consistent; both data sets capture high connectivity amongst the K2 outbreak strains, a distinct K3 sub-population, and divergent K4 infections^27^. The new infections appear to derive from across the different sub-populations.

Further spatial evaluations of the High-diversity SNP microhaplotype panel were undertaken to assess the accuracy of the panel in detecting imported *P. vivax* cases by determining an infection’s country of origin. Using the data from 21 countries with a minimum sample size of 4 from the available *P. vivax* global dataset, we determined the accuracy of the 494 SNPs within the microhaplotype panel in predicting country of origin using a recently developed Bi-Allele Likelihood (BALK) classifier^28^. The predictive accuracy of the 494 microhaplotype SNPs was compared to the 38-SNP Broad barcode and three recently identified *P. vivax* geographic barcodes selected specifically for determining country of origin (GEO33, GEO50 and GEO55 comprising 33, 50 and 55 SNPs respectively)^28^. The median Matthew’s correlation coefficient (MCC, ranging from -1 when prediction is 0% correct to 1 when prediction is 100% correct) of the 494 microhaplotype SNPs were greater than or equal to the 38-SNP Broad barcode, GEO33 and GEO50 in 21 (100%) countries (**Table 5, Figure 6**). The GEO55 panel exhibited higher MCCs than the microhaplotype panel in 3/21 (14%) countries (Cambodia, Indonesia and Vietnam) but lower values in 4/21 (19%) countries (Afghanistan, India, Myanmar and Thailand). The median MCCs of the microhaplotype panel exceeded 0.9 in all countries except Cambodia and Vietnam (19/21, 90%); the median MCCs in these countries were also <0.9 at the 38-SNP Broad barcode and the three GEO panels.

**Table 5.** Summary of median MCC scores from comparative evaluations of country prediction performance between the SNP panels. Comparisons were undertaken between the 494 SNPs in the High-diversity microhaplotype panel (MHAP-3_10), the 38-SNP Broad barcode (BR38), and the 33-, 50- and 55-SNP GEO panels (GEO33, GEO50 and GEO55 respectively). The median Mathews correlation coefficient (MCC) summary statistics are based on 500 repeats of the stratified 10-fold cross-validation using the Bi-Allele Likelihood (BALK) classifier in a set of *n*⍰=⍰799 biologically independent samples from 21 countries (each with n ≥4)^28^.

| Country | BR38 | GEO33 | GEO50 | GEO55 | MHAP-3_10 |
| --- | --- | --- | --- | --- | --- |
| Afghanistan | 0.835 | 0.850 | 0.908 | 0.889 | 0.946 |
| Bangladesh | 0.784 | 1.000 | 1.000 | 1.000 | 1.000 |
| Bhutan | 1.000 | 1.000 | 1.000 | 1.000 | 1.000 |
| Brazil | 0.892 | 0.892 | 1.000 | 1.000 | 1.000 |
| Cambodia | 0.520 | 0.756 | 0.823 | 0.874 | 0.866 |
| China | 0.864 | 1.000 | 1.000 | 1.000 | 1.000 |
| Colombia | 0.870 | 1.000 | 1.000 | 1.000 | 1.000 |
| Ethiopia | 0.923 | 0.933 | 1.000 | 1.000 | 1.000 |
| India | 0.794 | 0.864 | 0.892 | 0.864 | 1.000 |
| Indonesia | 0.913 | 0.939 | 0.985 | 1.000 | 0.985 |
| Iran | 1.000 | 1.000 | 0.892 | 1.000 | 1.000 |
| Madagascar | 1.000 | 1.000 | 1.000 | 1.000 | 1.000 |
| Malaysia | 0.784 | 0.933 | 0.933 | 0.933 | 0.933 |
| Mexico | 1.000 | 1.000 | 1.000 | 1.000 | 1.000 |
| Myanmar | 0.620 | 0.910 | 0.910 | 0.892 | 1.000 |
| Papua New Guinea | 0.344 | 0.812 | 1.000 | 1.000 | 1.000 |
| Peru | 0.817 | 0.940 | 1.000 | 1.000 | 1.000 |
| Philippines | 0.661 | 0.892 | 0.864 | 1.000 | 1.000 |
| Sudan | 1.000 | 1.000 | 1.000 | 1.000 | 1.000 |
| Thailand | 0.591 | 0.930 | 0.975 | 0.976 | 1.000 |
| Vietnam | 0.284 | 0.523 | 0.672 | 0.769 | 0.749 |
| **Pooled median** | **0.835** | **0.933** | **1.000** | **1.000** | **1.000** |
| **Pooled min** | **0.284** | **0.523** | **0.672** | **0.769** | **0.749** |
| **Pooled Q1** | **0.753** | **0.892** | **0.910** | **0.965** | **1.000** |

**Figure 6.**
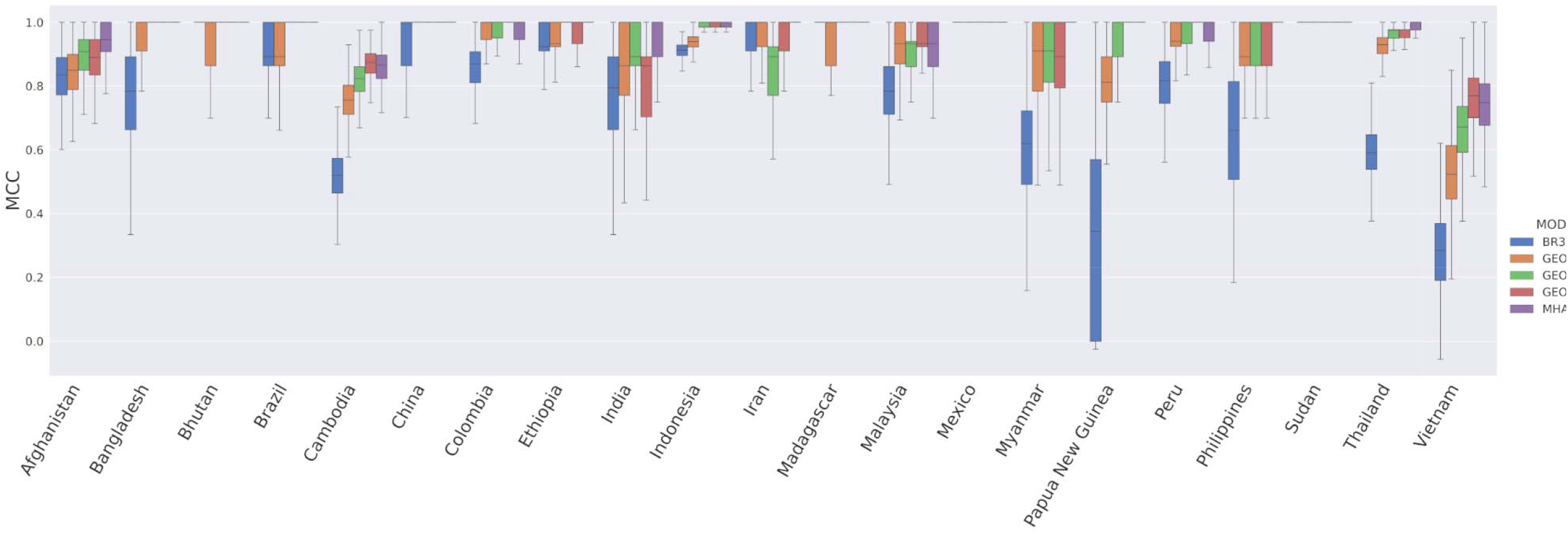
Comparison of country prediction performance between SNP panels. Comparisons were undertaken between the 494 SNPs in the High-diversity microhaplotype panel (MHAP-3_10), the 38-SNP Broad barcode (BR38), and the 33-, 50- and 55-SNP GEO panels (GEO33, GEO50 and GEO55 respectively). The boxplo present the Mathews correlation coefficient (MCC) scores from 500 repeats with stratified 10-fold cross validation for each SNP set using the Bi-Allele Likelihood classifier^28^. Country labels are provided on the y-axis. Each bar presents the median, interquartile range and minimum and maximum MCC for the given country and model. The analyses were based on *n*⍰=799 biologically independent samples from 21 countries (each with n ≥4 samples).

## Discussion

Our study provides the first description in silico of *P. vivax* microhaplotype panels which can be used to estimate “identity by descent” relatedness between pairs of acute and recurrent infection isolates, and thus help to discriminate between different causes of vivax malaria recurrence. A systematic genome-wide selection process was used to identify panels of 100 globally diverse microhaplotypes using an expansive *P. vivax* genome dataset. The utility of these panels was assessed against a pre-existing *P. vivax* SNP panel using both simulated and whole genome data. These relatively parsimonious panels have significant potential to improve the interpretation of clinical trials and surveillance data. Further details on the integration of microhaplotype data in the proposed use cases, and areas requiring further research and development are described herein.

A key requirement of the *P. vivax* microhaplotype panels was the ability to estimate IBD accurately between paired isolates and thereby help to classify the likely origin of vivax malaria recurrence (i.e., relapse, recrudescence or reinfection) in clinical trials. Our simulations showed that the microhaplotype panels yielded consistently higher accuracy in IBD estimation (across different IBD levels and in different populations) than the 42-SNP Broad barcode that is currently the most widely used SNP barcode for *P. vivax*. This is not surprising as the microhaplotypes have substantially greater genetic information content compared with the 38 evaluable Broad SNPs. We also demonstrated greater efficacy of IBD estimation when microhaplotypes were selected to meet specified diversity criteria (High-diversity SNP panel) relative to microhaplotypes with random SNP selections (any-SNP panel). In accordance with previous predictions^18^, our simulation-based results demonstrated error rates below 12% in the estimation of pairwise IBD in all populations tested using the high-diversity microhaplotype panel. The error rates were highest for the prediction of infections with 50% IBD. Further work is needed to understand how this might impact clinical predictions of individual treatment response or population-level drug efficacy both in the context of descriptive data analyses and mathematical models^29^. In descriptive analyses, data summaries (e.g., allelic homology observations or relatedness estimates) and a set of rules (e.g., relatedness values greater than 25% are suggestive of relapse) are used to classify recurrences categorically. Rules-based classification is not the same as estimating the probability of relapse given the data under a statistical model (as in Taylor et al^29^, which also accounts for the added complexity associated with multiclonal infections). Nevertheless, improved data informativeness for IBD estimation likely translates into improved model-based probabilistic classification performance, at least in the case of the model of Taylor, because that model machinery includes an intermediary that evaluates the probability of the data given IBD partitions compatible with networks of sibling, clonal and stranger parasites^29^.

It should be acknowledged that panel evaluation based on simulated data was done in a highly idealised manner that captures panel performance in its most favourable light. Data were simulated under the same model used to estimate relatedness, such that the model was perfectly specified in relation to the simulated data. Real data are generated by an ancestral process that the model does not capture, i.e., the model is miss-specified in relation to real data. The impact of miss-specified allele frequencies, miss-classified multiclonal infections, etc. was not evaluated. In addition, it should be acknowledged that genotyping errors and failures (missing data) were not included in our simulations. However, we anticipate that genotyping failures should be infrequent with NGS-based amplicon sequencing methods, where each locus is typically covered by hundreds of reads^19^.

Data from pairs of *P. vivax* isolates, collected from recurrent infections in clinical trials, confirmed the ability of the high diversity microhaplotype panel to estimate pairwise IBD. High correlations were observed between microhaplotype and genome wide IBD estimates in a set of 14 paired *P. vivax* isolates from the same patient before and after treatment. Using our assigned IBD thresholds, only one of 14 pairs of infections had mismatching classifications of ‘stranger’ versus ‘distant relative’, which are both likely to reflect either reinfections or relapses. However, the available genomic data did not comprise any infection pairs with IBD estimates around 50% (i.e., siblings), which our simulations predicted to be the most difficult relationships to determine accurately. The number of genomic pairs was limited owing to the difficulty in obtaining enough DNA and thus high-quality sequence data from recurrent infections, which typically exhibit low parasite densities. This is a major incentive for using targeted genotyping approaches. Nonetheless, we demonstrated high correlations between the microhaplotype and genome wide IBD estimates in the assessments of day 0 samples, where a range of IBD relationships were observed.

In some scenarios, genetic data alone will not be informative of the likely origin of a recurrence. A single mosquito inoculation may carry clonal parasites or related parasites generated through recombination of heterologous clones. If a human host carries hypnozoites from a single inoculation, the relapsing parasites will therefore either be homologous or related (meiotic siblings) to the incident infection. In situations where the relapses are homologous, they will be genetically indistinguishable from recrudescence. If a host carries hypnozoites from one or more previous mosquito inoculations, relapsing parasites are liable to be unrelated/heterologous to the incident infection, and thus genetically indistinguishable from reinfections. In our study, a high proportion of infection pairs (79%) had homologous genomes, although the majority (86%) of pairs derived from low seasonal transmission settings in Malaysian Borneo, southern Vietnam, Cambodia and western Thailand. A therapeutic efficacy study undertaken in Peru also identified a moderately high proportion of genetically homologous recurrences (52%, 12/23)^12^. The proportion of relapses amongst the recurrences in our and the Peruvian study is uncertain. In a therapeutic efficacy study undertaken in Cambodia, 20 patients were relocated to a malaria-free area excluding the possibility of reinfection; the authors were able to define confidently 5 recurrences as relapses and demonstrated that 4 (80%) of these were related to the initial infection^30^. As more comprehensive data become available, a clearer picture of the epidemiology of recurrent *P. vivax* infections will emerge. Even in areas where the prevalence of related recurrences is low, mathematical modelling approaches that combine genetic data with time-to-event data will help to resolve the probable cause of recurrence^29^.

Another key requirement of the microhaplotype panel was the ability to capture spatial *P. vivax* transmission dynamics. Strategies to contain *P. vivax* effectively will be assisted by a more comprehensive understanding of the major routes of infection spread within and across borders. The SNP-based data from our microhaplotype panel displayed clear geographic trends and high accuracy in predicting the country of origin, suggesting utility in detecting and mapping imported *P. vivax* cases. Although the high-diversity microhaplotype panel was not intentionally selected for country prediction, we captured 494 high diversity bi-allelic SNPs with 100 microhaplotypes and these highly informative genetic data enabled high performance country prediction compared to recently described geographic marker panels (GEO33, GEO50 and GEO55) that were selected specifically to differentiate *P. vivax* populations by country^28^. The accuracy in pairwise IBD estimation using the microhaplotype data also demonstrates a unique potential for tracking infection spread at micro-epidemiological spatial resolution, to inform the dispersal of infections within and between communities. For example, the microhaplotype data from Malaysia effectively captured a previously described clonal expansion, as well as more subtle population structure reflecting different foci of infection. The spatial analyses conducted here used biallelic SNP data at the microhaplotypes; whilst the high density of SNPs (n=494) provided rich genetic information, even greater information content can be achieved using multiallelic microhaplotypes once new software to deal with these complex datasets becomes available.

Some global regions are not well represented in the Pv4 genomic dataset that were used for marker selection, such as the Indian subcontinent, Central and South America. It is therefore unclear how well the microhaplotypes described here will capture IBD in these regions. However, the informatics pipeline that was established for the microhaplotype selection can be applied readily to update the panel as needed once additional whole genome data become available from new geographical regions. The pipeline can also be used to select country-or region-specific panels where needed.

Information on within-host infection complexity is important to capture epidemiologically relevant transmission dynamics. We observed high concordance in the proportion of polyclonal infections captured by microhaplotype-based COI and genome-wide *F*_*WS*_ measures when thresholds of COI=1 and *F*_*WS*_ ≥0.95 were applied. Only a few of infections displayed differences in classification of polyclonality between the microhaplotype and genomic datasets. In interpreting these differences, it should be acknowledged that the 0.95 *F*_*WS*_ threshold is only a guideline, and that population distributions of within-host diversity generally reflect a continuum, not discrete clusters of monoclonal and polyclonal infections.

Within-host diversity at SNP barcodes also follows a continuum, but this is less marked owing to the lower genetic resolution. When monoclonal thresholds were only applied to the COI data, the *F*_*WS*_ demonstrated significantly larger values in the COI=1 (monoclonal) vs COI>1 (polyclonal) infection group in all geographic regions investigated, highlighting consistency between the two data sets. Further work is needed to determine how to phase microhaplotype profiles in highly complex infections where any number of clones may be present in varying proportions, but tools such as the *Dcifer* software provide an important step forward^31^. Indeed, phasing of polyclonal malaria infections is not a challenge unique to microhaplotypes.

The *P. vivax* genome has an abundance of globally diverse microhaplotype regions that can effectively capture information on infection lineages and spatial connectivity, overcoming the previous requirement to generate genomic data in samples that are often notoriously difficult to sequence. With targeted, deep sequencing platforms, these markers have great potential to inform on the complex diversity within individual infections and associated insights on transmission. The establishment of targeted microhaplotype genotyping tools for *P. vivax* will transform the assessment of clinical surveys in this species, enhance knowledge of relapse biology, and greatly improve surveillance of infections.

## Methods

### Data preparation

The initial dataset comprised 1,816 samples from 17 countries derived from the *P. vivax* community study (Pv4.0) in the vivax and Malaria Genomic Epidemiology Networks (vivaxGEN and MalariaGEN)^20^, as well as previously published external studies^32-34^. Of the 1,895 samples described in Pv4.0 data note, the 1,816 samples reflect data for which we had access prior to the open release. All genomic datasets were generated using Illumina short-read sequencing platforms. Sequence alignment, SNP discovery and variant calling, population assignments, and *F*ws (within-sample F statistic) calculations for within-host allele infection complexity were undertaken using previously described methods within the MalariaGEN framework, producing a dataset referred to as *P. vivax* Genome Variation Project release 4.0 (Pv4.0). The *F*_*WS*_ estimates the fixation of alleles within each infection relative to the diversity observed in the total population on a scale from 0 to 1 and were provided as part of the Pv4 dataset^20,35^. Using a threshold of *F*_WS_ ≥0.95 as a proxy to identify a monoclonal infection, all polyclonal infections were excluded from subsequent analysis. High-quality samples were selected using a threshold of ≥50% core genome, notated by the “analysis-set” flag in the MalariaGEN Pv4 dataset. The patient metadata provided by the contributing VivaxGEN partner studies was used to identify independent isolates for marker selection, and recurrent isolates for downstream evaluation of selected marker sites. The MalariaGEN curated metadata was used to define 7 regional-level geographic assignments with the following categories: Africa (AF), South and Central America (SAM), West Southeast Asia (WSEA), East Southeast Asia (ESEA), Maritime Southeast Asia (MSEA), Oceania (OCE) and West Asia (WAS). The regional classifications were based on genomic clustering patterns in the Pv4.0 data notated as “analysis population” flag.

### Marker selection

We tailored our marker selection to the Illumina amplicon-based sequencing workflow as this methodology is widely used in the malaria community and has demonstrated utility and feasibility for malaria molecular surveillance in low- and middle-income countries^17^. The maximum amplicon size for Illumina amplicon-based sequencing is 200 bp, which set the criteria for the maximum length of each microhaplotype. The target number of microhaplotypes was set to 100 based on a mathematical modelling study, which demonstrated that under idealised settings (i.e., estimating relatedness from data simulated under the model used for estimation), IBD estimates with low root mean squared error (RMSE<0.1) between monoclonal malaria parasites with relatedness equal to 0.5 (relatedness with the highest RMSE) are obtainable using ∼100 polyallelic markers^18^. Diminishing returns in RMSE reduction were observed above 100 polyallelic markers, highlighting this target number as a pragmatic balance of IBD accuracy against the economic cost of primers^18^. Panels of microhaplotypes (schematic **Figure 1a**) were subsequently selected to optimise marker selection with the previously identified criteria using a three-phase approach as described in **Figure 1b** with respect to sample, variant and window selection. In the first phase, samples were subset from the MalariaGEN Pv4.0 dataset that consisted of high-quality samples as identified in Pv4 as being independent, 50% of the genome callable, and likely monoclonal with *F*ws ≥0.95. In the second phase, candidate SNPs were identified by filtering the high-quality biallelic SNPs (filter pass), with minor allele frequency (MAF) <0.1 and genotype failure rates >0.1. Finally, candidate microhaplotype windows were identified by scanning the core genome (excluding hypervariable regions) in coding regions with 50 bp overlapping, 200 bp windows to identify amplicon-sized segments containing at least one candidate SNP, producing what we define as the heterozygome map of the accessible regions of the *P. vivax* genome (**Figure 1c**). Once the windows were identified, they were further filtered to a minimum heterozygosity of 0.5 for a total of 5,460 windows. Panels of 100 microhaplotypes were then selected from the *P. vivax* heterozygome by selecting only windows that had between 3-10 SNPs, and approximately evenly distanced spacing across 14 chromosomes. The “Random mhaps” panel had 100 markers chosen irrespectively of diversity (with a minimum of 0.5 heterozygosity) and the “High-diversity” panel had each of its markers chosen with the highest possible heterozygosity in a given genomic region (Figure 1d). Measures of MAF and heterozygosity were calculated using the scikitallel package.

### Population genetic evaluation of the microhaplotype panel

To evaluate the efficacy of the selected microhaplotype panel relative to the Broad *P. vivax* barcode as a global barcode, population diversity was assessed at each panel (microhaplotype and Broad) for each of the 7 geographic regions using measures of effective cardinality and heterozygosity computed with the R-based *paneljudge* package (https://github.com/aimeertaylor/paneljudge). Analyses were conducted on the high-quality monoclonal samples using one of the two alleles selected at random to reconstruct microhaplotypes at heterozygote genotype positions using the “haploidify” function in Scikit-allel. Measures of within-host diversity were also generated using *THE REAL McCOIL* package on the biallelic SNPs in the High-diversity microhaplotype panel and correlated against the genome-wide *F*_*WS*_ estimates of infection complexity^35,36^.

The potential of the biallelic SNPs within the microhaplotype panel in capturing spatial patterns of transmission was assessed using Principal Coordinate Analysis (PCoA) to illustrate the regional level geographic clustering patterns. PCoA analysis was performed on the same high-quality monoclonal samples (n=615) by conducting PCA measures using *sklearn*.*decomposition PCA* method of scikit-learn package version 1.2.0 on a microhaplotype-based pairwise distance matrix calculated from the proportion of non-identical microphalotype alleles across all positions. Spatial clustering patterns were also assessed using IBD-based measures of differentiation implemented with the hmmIBD package as described below and plotted using the R-based *igraph* package (https://igraph.org).

A Bi-allele Likelihood (BALK) classifier was used to predict the country of origin of *P. vivax* isolates using SNP data (not multi-allelic microhaplotypes) at biallelic SNPs in the High-diversity SNP microhaplotype panel, Broad *P. vivax* barcode, and three recently identified *P. vivax* geographic barcodes (GEO33, GEO50 and GEO55)^21,28^. Details of the BALK classifier can be found in the original paper^28^. Country prediction performance was assessed using data from 799 of the 922 high-quality samples in the available Pv4.0 genomic dataset. The 799 sample-set corresponds with the sample set used in the original paper describing the BALK classifier^28^. Briefly, the sample set was derived by filtering samples to include a single representative of samples with near-identical genomes (defined as pairs with genetic distance <0.001), subjecting to iterative data quality filtering to obtain the best representative number of samples and informative SNPs without any missing genotype by removing samples with higher missingness iteratively, and then removing samples that appeared to be imported based on genome-wide data clustering patterns. A total of 21 countries had ≥4 samples. The comparative predictive performance of the SNP panels was evaluated using a stratified 10-fold cross-validation with 500 repeats, reporting the Matthews Correlation Coefficient (MCC).

### Evaluation of IBD estimation with the microhaplotype panel

The 2 selected microhaplotype panels (Random microhaplotype panel and High-diversity microhaplotype panel), as well as the 42-SNP Broad barcode panel, were evaluated with *paneljudge* package for their ability to estimate relatedness (IBD) using a range of simulations to compute the uncertainty (error rate) in estimations of relatedness (*r*) ranging from 0 (strangers) to 1 (identical) under varying allele frequency distributions in different geographic regions. Details of the equations, assumptions, and parameter options in the *paneljudge* package can be found on GitHub (https://github.com/aimeertaylor/paneljudge). Briefly, for each of a range of data-generating *r* values (*r* = 0, 0.25, 0.5, 0.75 and 1.0), data were simulated on a pair of haploid genotypes, generating 100 haploid genotype pairs. Estimates of *r* 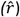, switch-rate parameter 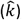, the 95% confidence intervals (CIs) around the estimates 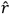 and 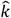, and root mean square errors (RMSE) were then computed. The switch-rate parameter modifies rate of switching between latent IBD and non-IBD states in the hidden Markov model by multiplying the probability of a crossover per base pair (default value 7.4e-7 as per ^37^) and the inter-marker distance in base pairs. The accuracy of a given marker panel in estimating *r* was measured by the CI width and RMSE, ranging from ∼0 (near perfect informativeness) to 1 (uninformative). The CI width and RMSE of 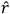 compared to the data-generating *r* never reach 0 as a genome of finite length may have many realised relatednesses compatible with a given probability of IBD. This means that there will always be some variance in realised relatedness around *r*. For comparative value, *paneljudge* simulations were run on a panel of 100 single SNP markers (i.e., biallelic markers) and for a set of 38 assayable biallelic SNPs from the 42-SNP Broad barcode that has been widely used by the *P. vivax* surveillance community^21,38,39^. The 100-SNP biallelic panel was composed by selecting the highest diversity SNP in each microhaplotype.

In addition to simulation-based data, the efficacy of the selected microhaplotype panel in estimating IBD was assessed relative to ‘gold-standard’ genome-wide data in pairs of primary (day 0) and recurrent *P. vivax* infections with high-quality genomic data (≥50% genome covered by at least 5 reads) which were monoclonal infections (*F*_*WS*_ ≥0.95). IBD estimates between infection pairs were generated using the *hmmIBD* version 3.0 software^40^. As the version 3 software does not enable adjustments in the genotyping error rate for polyallelic markers (which may reduce artificially the IBD estimates between infections) the microhaplotype data were run as biallelic SNP markers rather than polyallelic markers. Sample pairs were run in geographically defined batches using allele frequency estimates based on the regional estimates from the baseline samples (high-quality, monoclonal, independent infection) using default parameters. Genome-wide and microhaplotype-based recurrence classifications were assigned using the following infection pair IBD thresholds; clone (pairwise IBD ≥0.95), close relative (0.25≤ IBD <0.95), distant relative (0.05≤ IBD <0.25), stranger (IBD <0.05). The concordance between the microhaplotype and genome wide IBD estimates was evaluated by the correlation coefficient between the datasets, and the proportion of recurrence classification mismatches.

## Data Availability

All data produced in the present study are available upon reasonable request to the authors

## Ethics

As detailed in a corresponding data note for the Pv4.0 data set, all isolates were collected in the framework of studies with ethical approval from the local research ethics boards, and with written, informed consent provided by patients or guardians for individuals of age 18 or younger^20^.

## Data availability

Sequencing data from these samples has been made publicly available in the European Nucleotide Archive (ENA), with details provided in a data note describing the MalariaGEN Pv4.0 data set^20^.

## Code availability

The custom, in-house scripts used to select the microhaplotype panels are available from https://github.com/svsiegel/vivax-mhaps.

## Acknowledgements

We thank the patients who contributed their samples to the study, and the health workers and field teams who assisted with the sample collections. Genome sequencing was undertaken by the Wellcome Sanger Institute and we thank the staff of the Wellcome Sanger Institute Sample Logistics, Sequencing, and Informatics facilities for their contribution.

## Supplementary Material

**Supplementary Table 1. Microhaplotype panel marker selection information**.

**Supplementary Table 2.** Properties of the three most diverse markers in each geographic region for the High-diversity microhaplotype panel. *The PvP01_05_v1:1369384:7,52,99,114,121,126,136,164 microhaplotype was one of the top three most diverse markers in all seven geographic regions.

| Region | Chromosome: coordinate: SNP position | Effective cardinality | Heterozygosity |
| --- | --- | --- | --- |
| AF | PvP01_14_v1:3009931:19,27,72,78,81,89,119,139,150 | 18.94 | 0.95 |
| AF | PvP01_07_v1:78144:4,11,28,79,127,130,136,156 | 10.25 | 0.9 |
| AF | *PvP01_05_v1:1369384:7,52,99,114,121,126,136,164 | 9.62 | 0.9 |
| ESEA | *PvP01_05_v1:1369384:7,52,99,114,121,126,136,164 | 21.28 | 0.95 |
| ESEA | PvP01_01_v1:772094:42,60,64,65,117 | 12.97 | 0.92 |
| ESEA | PvP01_07_v1:78144:4,11,28,79,127,130,136,156 | 10.69 | 0.91 |
| MSEA | *PvP01_05_v1:1369384:7,52,99,114,121,126,136,164 | 23.91 | 0.96 |
| MSEA | PvP01_11_v1:1423352:18,64,79,126,137,160,167 | 15.24 | 0.93 |
| MSEA | PvP01_14_v1:2261605:80,96,114,135,189 | 11.9 | 0.92 |
| OCE | *PvP01_05_v1:1369384:7,52,99,114,121,126,136,164 | 26.05 | 0.96 |
| OCE | PvP01_11_v1:1423352:18,64,79,126,137,160,167 | 17.93 | 0.94 |
| OCE | PvP01_06_v1:286251:5,6,28,75,168,179,184 | 13.64 | 0.93 |
| SAM | *PvP01_05_v1:1369384:7,52,99,114,121,126,136,164 | 27.16 | 0.96 |
| SAM | PvP01_11_v1:1423352:18,64,79,126,137,160,167 | 19.94 | 0.95 |
| SAM | PvP01_06_v1:286251:5,6,28,75,168,179,184 | 16.57 | 0.94 |
| WAS | *PvP01_05_v1:1369384:7,52,99,114,121,126,136,164 | 11.72 | 0.91 |
| WAS | PvP01_06_v1:983158:17,19,24,35,117,136,175 | 10.74 | 0.91 |
| WAS | PvP01_06_v1:286251:5,6,28,75,168,179,184 | 8.54 | 0.88 |
| WSEA | *PvP01_05_v1:1369384:7,52,99,114,121,126,136,164 | 36.9 | 0.97 |
| WSEA | PvP01_11_v1:1423352:18,64,79,126,137,160,167 | 24.2 | 0.96 |
| WSEA | PvP01_06_v1:286251:5,6,28,75,168,179,184 | 15.39 | 0.94 |

**Supplementary Figure 1.**
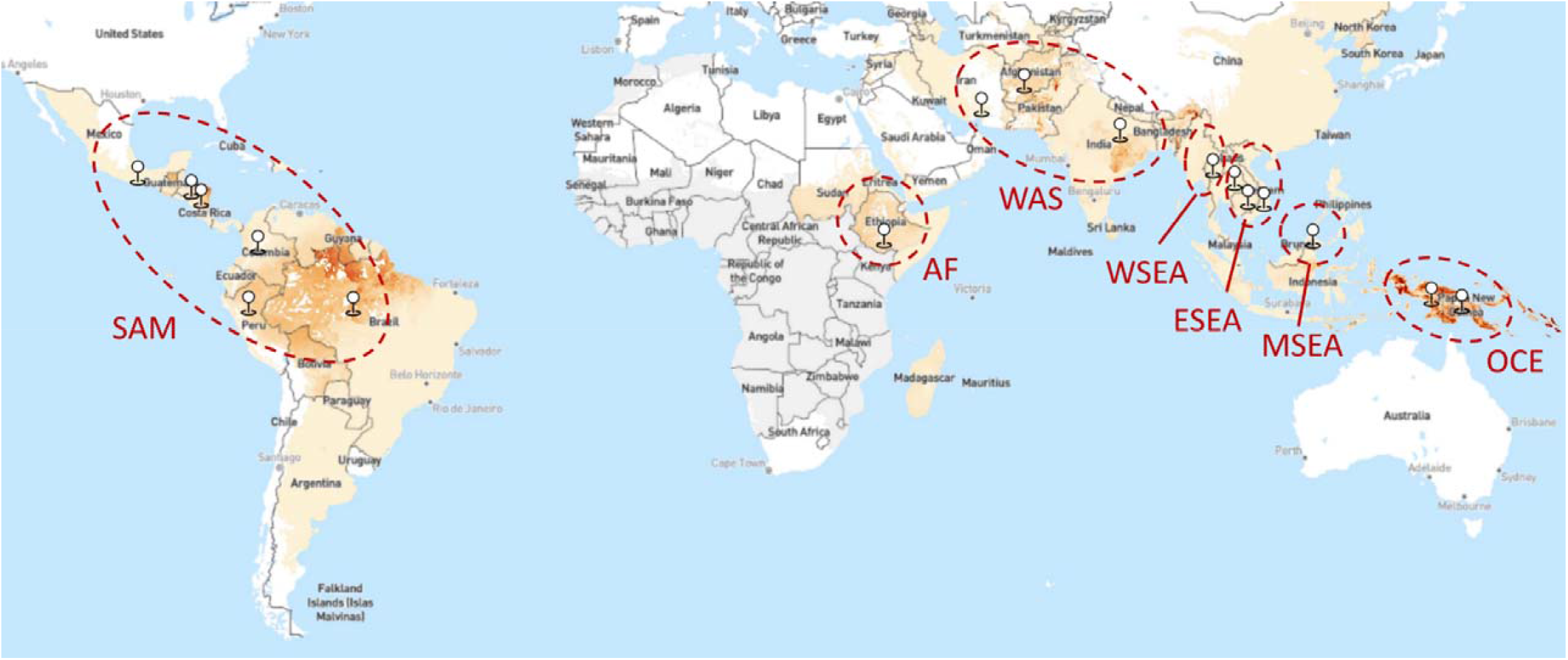
*P. vivax* incidence map illustrating regional country groupings. The baseline *P. vivax* incidence map was derived from the Malaria Atlas Proj (MAP) and presents the number of newly diagnosed *P. vivax* cases per 1,000 population in 2020^41^. The labelled, dashed red lines indicate the boundaries of the geographic regions included in the identity by descent (IBD) analyses: SAM (South America), AF (Africa), WAS (West Asia), WSEA (West Southeast Asia), ESEA (East Southeast Asia), MSEA (Maritime Southeast Asia) and OCE (Oceania). Pinpoints indicate the countries included in each regional grouping.

**Supplementary Figure 2.**
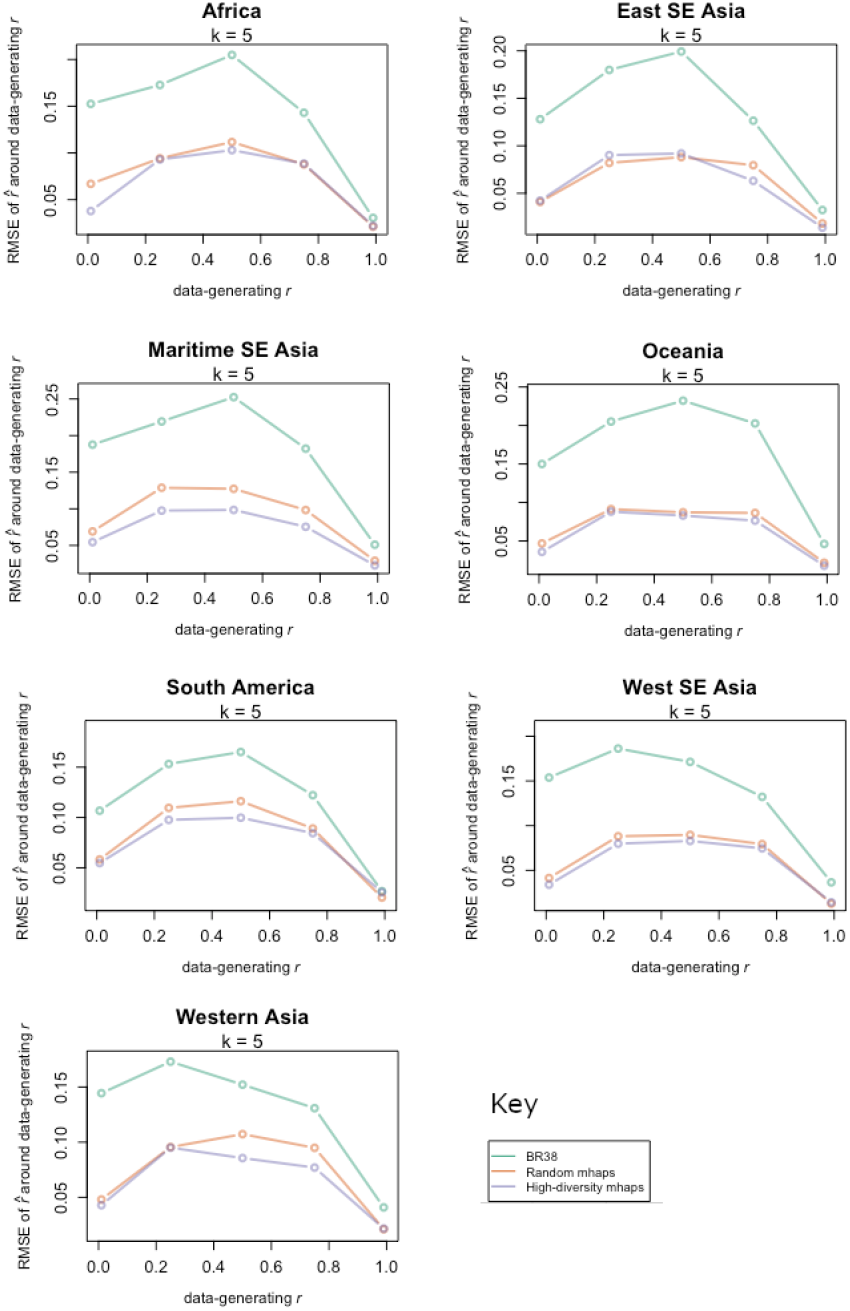
Comparative accuracy in relatedness prediction at Random and High-diversity microhaplotype panels, and the 38-SNP Broad barcode. Root mean square error (RMSE) of relatedness estimates based on data simulated using various data-generating relatedness and switch rate parameters, *r* and *k*, respectively. Data are presented on 3 marker panels: High-diversity SNP microhaplotype panel, Random-SNP microhaplotype panel and 38 Broad barcode biallelic SNPs. Panel comparisons are presented by geographic region; AF (Africa), ESEA (East Southeast Asia), MSEA (Maritime Southeast Asia), OCE (Oceania), SAM (South America), WAS (West Asia) and WSEA (West Southeast Asia).

**Supplementary Figure 3.**
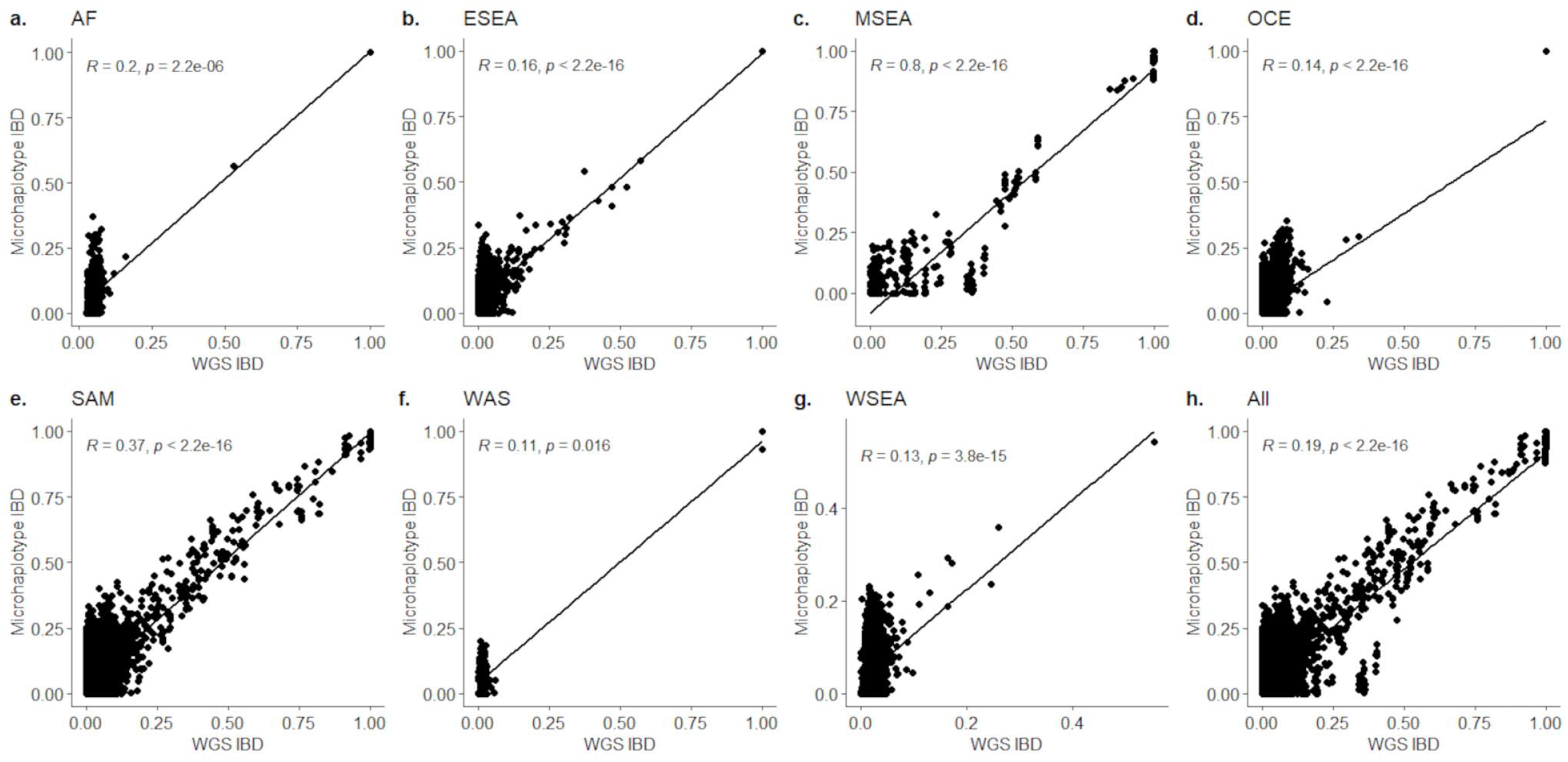
Correlations between microhaplotype and genomic estimates of IBD in the regional datasets. Panels a) to g) represent Africa (AF), East Southeast Asia (ESEA), Maritime Southeast Asia (MSEA), Oceania (OCE), South America (SAM), West Asia (WAS), and West Southeast Asia (WSEA). The microhaplotype and whole genome sequence (WGS) IBD estimates reflect pairwise estimates at the High-diversity SNP microhaplotype panel and a set of 898,448 genome-wide SNPs calculations were performed using *hmmIBD* on the 615 monoclonal sample set. Correlations were assessed with Spearman’s rho statistic (using a paired test) and presented with the associated p-value. At an alpha of 0.05, significantly positive correlations were observed in all regions.

**Supplementary Figure 4.**
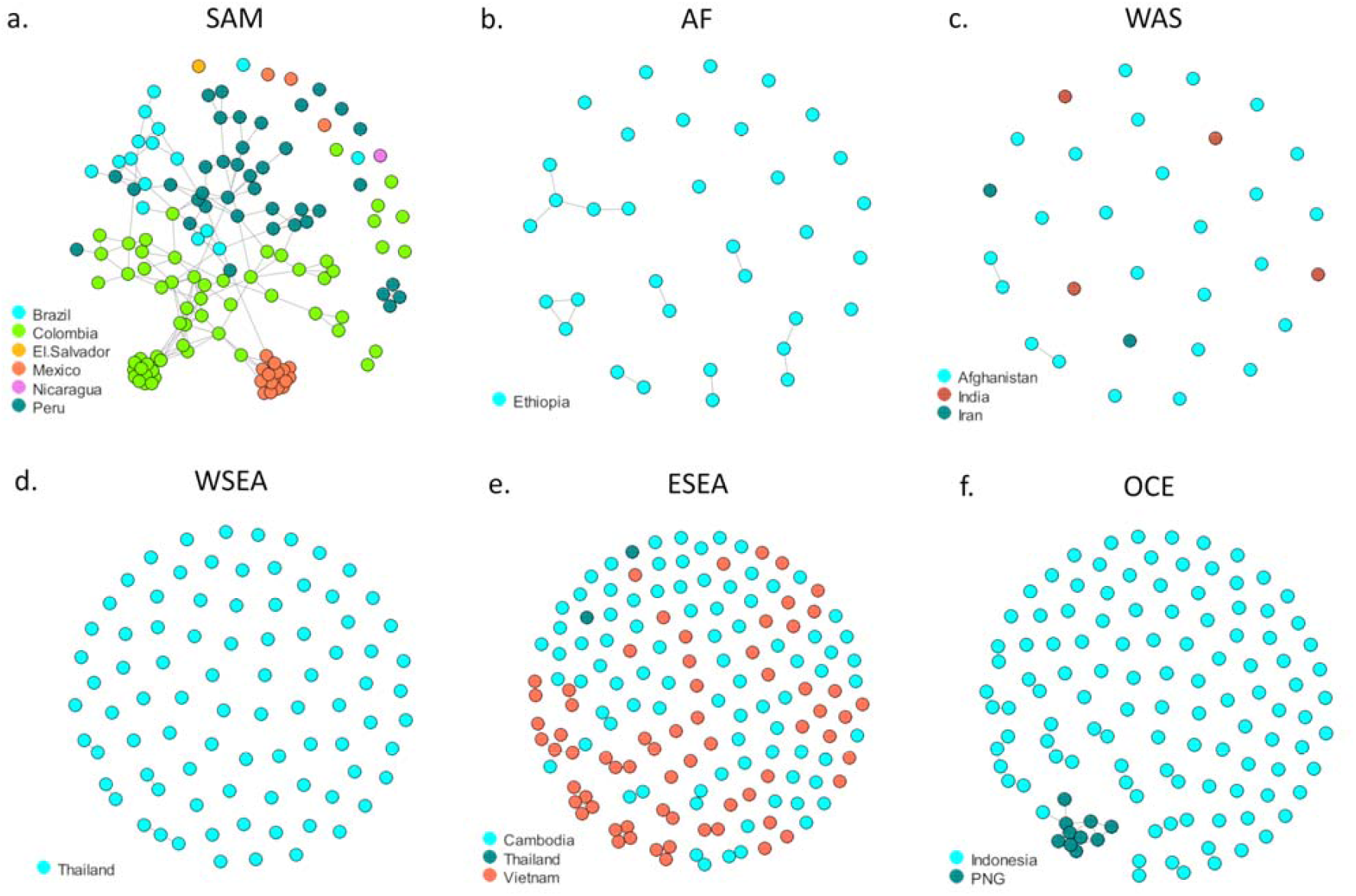
Regional microhaplotype-based infection networks. Panels a) to e) represent South America (SAM), Africa (AF), West Asia (WAS), West Southeast Asia (WSEA), East Southeast Asia (ESEA) and Oceania (OCE). The networks were generated from the High-diversity SNP microhaplotype panel in the 615 monoclonal sample set. The isolates from Maritime Southeast Asia (MSEA) were mostly from Malaysia (57/59) and are represented in Figure 5. Each circle reflects an infection, colour-coded by country, and line lengths reflect relatedness (shorter lines reflect greater relatedness) at a connectivity threshold of minimum identity by descent (IBD) 0.25 (half-siblings or greater relatedness). Each circle reflects an infection, with colour-coding by country.

