## Supplementary figures and images for "Lineage-informative microhaplotypes for spatio-temporal surveillance of *Plasmodium vivax* malaria parasites"

### Supplemental Figure 1

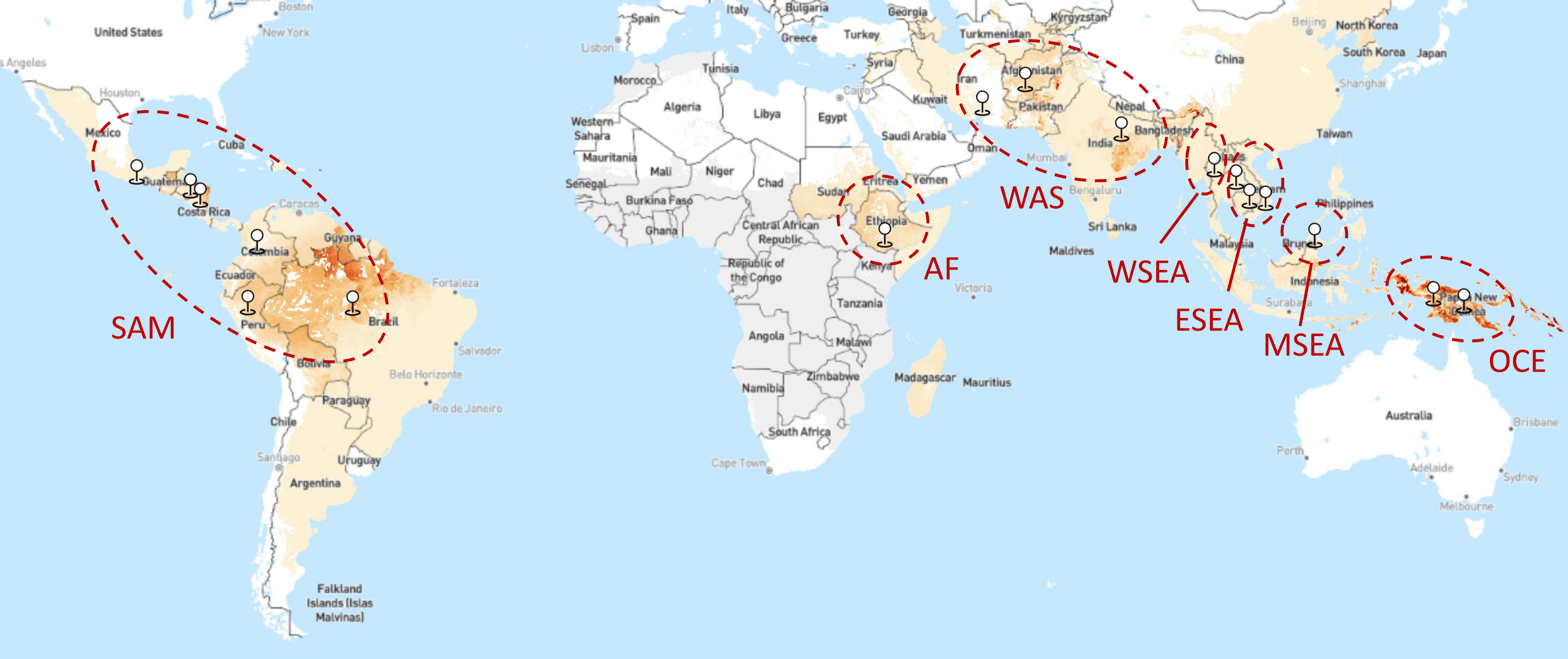

### Supplemental Figure 2

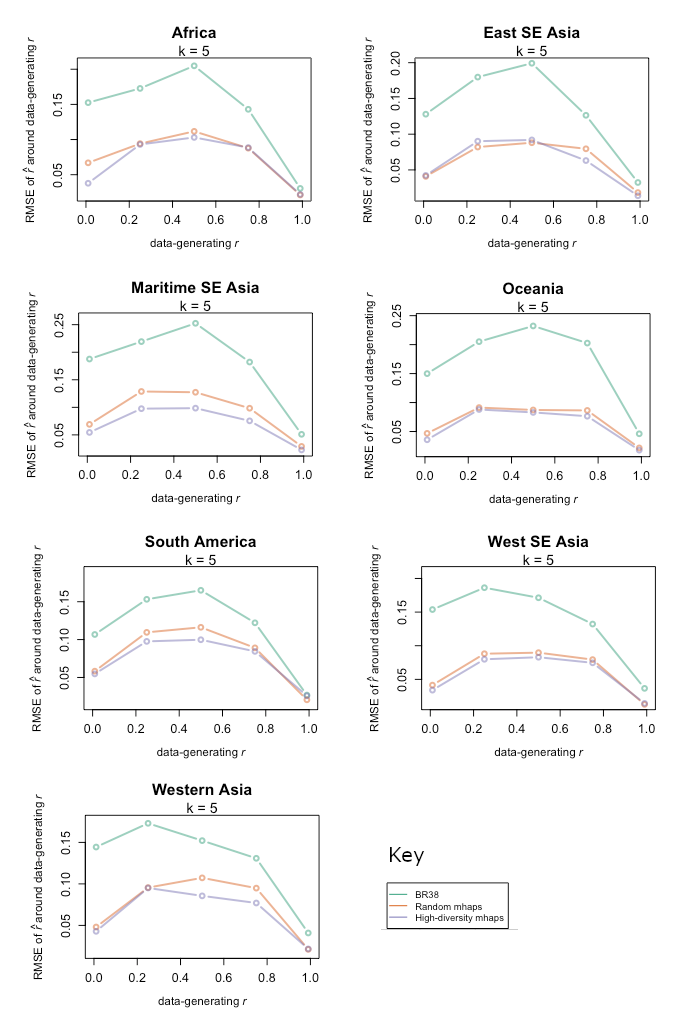

### Supplemental Figure 3

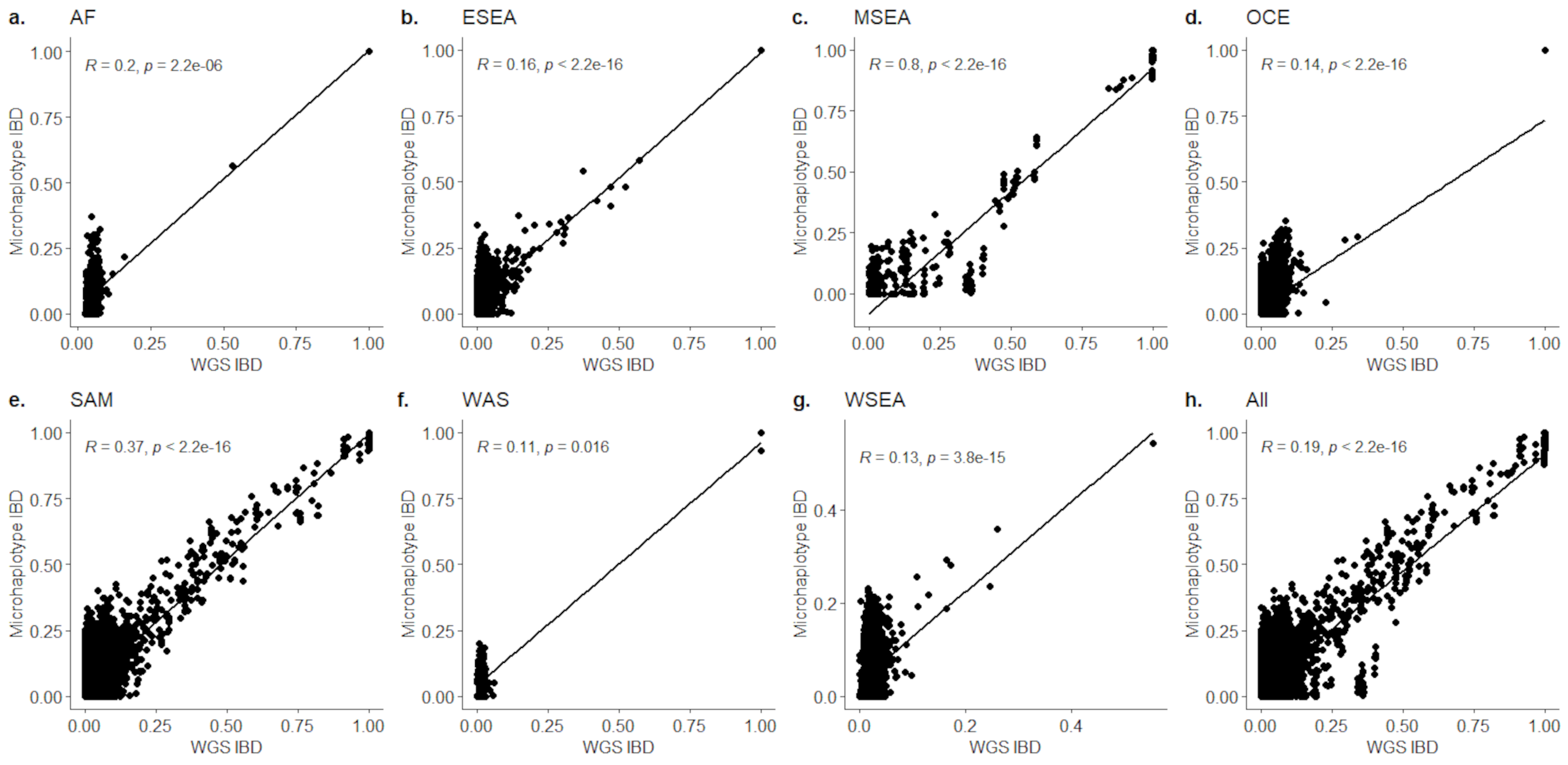

### Supplemental Figure 4

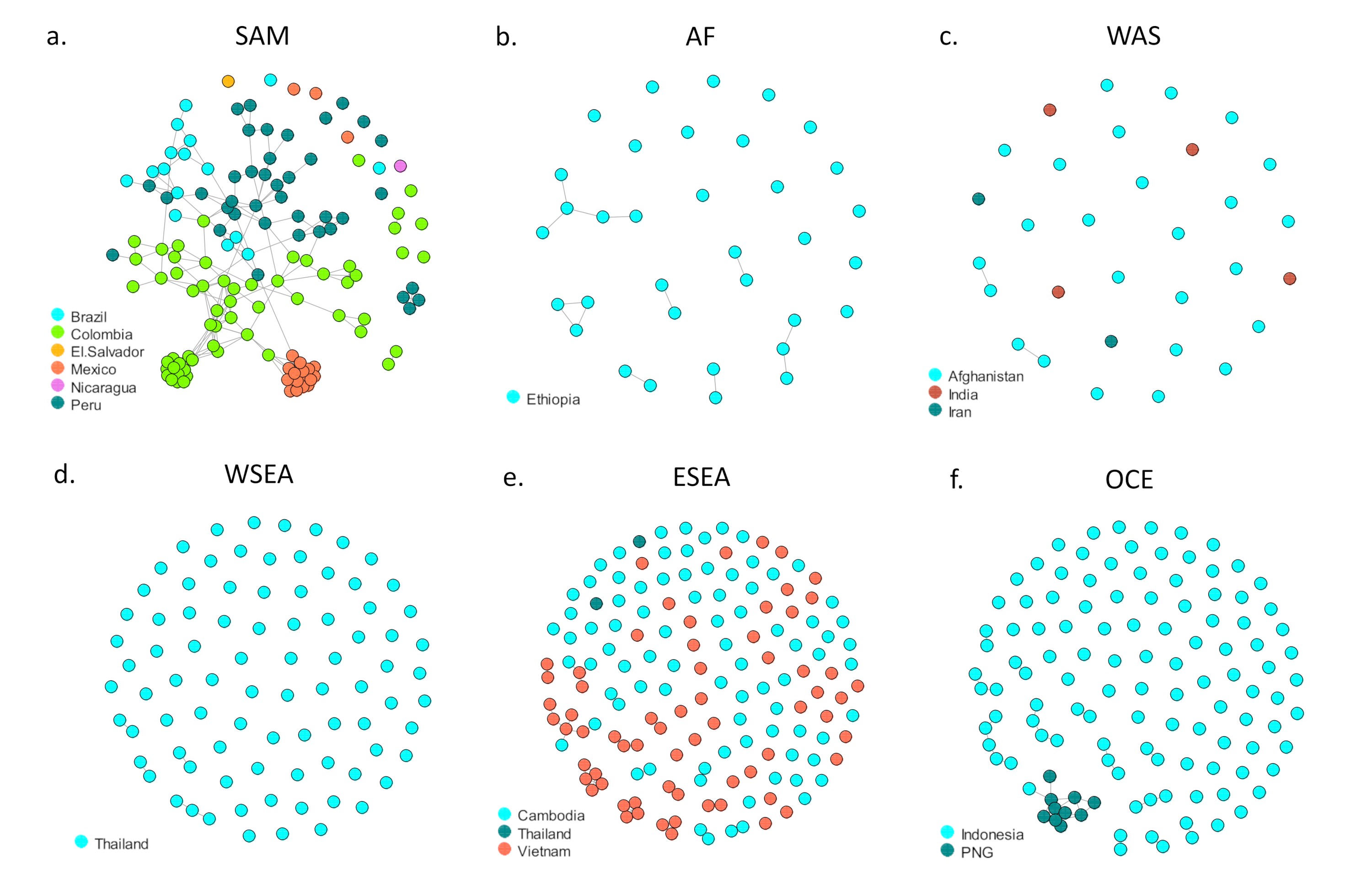
